# Multi-ancestry analysis of 791K whole genomes reveals the genetic, geographic, and phenotypic correlates of somatic passenger mutations in blood

**DOI:** 10.64898/2026.08.21.26361040

**Authors:** Prarthana Sanjeeva Reddy, Karen N. Conneely, Joshua S. Weinstock

## Abstract

Clonal hematopoiesis (CH), an aging-related expansion of hematopoietic stem cell (HSC) clones, is associated with hematologic malignancy, cardiovascular disease, and mortality. Most clonal expansions, however, occur in the absence of known driver mutations. Passenger mutations reveal positive selection in HSCs and provide a quantitative, driver-agnostic phenotype that increases statistical power over dichotomized driver-based definitions. We used somatic passenger mutation burden as a quantitative phenotype to map the genetic, phenotypic, and geographic correlates of CH across 791,067 blood whole-genome sequences from UK Biobank (UKB) and the All of Us Research Program (AoU). Multi-ancestry meta-analysis of genome-wide association studies in both cohorts identified 81 loci associated with passenger mutation burden, including 42 novel loci. Rare-variant analyses additionally implicated *MBD2*, *PRKACB*, *PUF60*, and related epigenetic and transcriptional regulators of clonal fitness. Together, common and rare germline associations converged with canonical CH drivers on shared pathways regulating DNA methylation, chromatin, RNA splicing, and genome maintenance. Sex and ancestry stratified analyses revealed the shared and population-specific determinants of CH, including loci undetected in the pooled analysis. Associated variants were concentrated in regulatory elements active in hematopoietic stem and progenitor cells, linking germline associations to relevant cell states and lineages. Phenome-wide analyses revealed distinct consequences of inherited CH liability and observed passenger burden, with passenger burden strongly associated with incident hematologic malignancy and mortality and germline risk showing additional nonhematologic associations. Finally, spatial modelling of passenger mutation burden across U.S. regions revealed persistent geographic heterogeneity incompletely explained by income, air quality, or chemotherapy prevalence, pointing to additional unmeasured environmental exposures. Overall, through parallel analyses of two population-scale biobanks, we characterize the multi-ancestry genetic architecture of CH and reveal convergence of germline and somatic variation on shared pathways governing clonal expansion in aging blood.

## Introduction

Clonal hematopoiesis (CH) is the age-associated expansion of hematopoietic stem (HSC) clones that have acquired somatic mutations conferring a fitness advantage ^1^. These clones compete throughout the human lifespan, and our blood bears the genetic scars of this competition later in life. CH is common in aged individuals, with recent prevalence estimates of 10%–20% in those aged over 70 years ^2,3^ and is linked to elevated risks of hematologic malignancy, atherosclerotic cardiovascular disease, heart failure, chronic kidney disease, and all-cause mortality ^3–7^.

CH is commonly defined in practice by the detection of driver mutations^8–12^. However, through whole-genome sequencing of single-cell-derived colonies of HSCs, recent reports have shown that most clonal expansions in aging blood occur in the absence of canonical driver mutations ^2^. Concordantly, the burden of passenger mutations has been shown to reveal pervasive unexplained positive selection in HSCs^13^. These observations indicate that our understanding of the etiology of CH remains incomplete.

The shadow of positive selection includes passenger mutations – somatic mutations that do not alter cell fitness. Through phenotyping of these hitchhikers, previous reports ^14–17^ have examined CH and its genetic and phenotypic correlates without ascertainment of specific genetic lesions. In contrast to driver mutations, passenger mutation abundance is quantitative, and thus allows more granular analysis than dichotomized driver definitions.

Recent papers have shown that germline variation associates with CH. The germline background of CH in an individual shapes one’s susceptibility through 1) modulating the rate at which HSPCs acquire somatic mutations and/or 2) altering the selective fitness advantage that drives clonal expansion ^14–16,18^. In this work, we demonstrate that germline variation associated with a driver-agnostic quantitative passenger burden phenotype not only recapitulates canonical CH loci such as *TERT* and *TCL1A* but also reveals dozens of novel loci, including epigenetic regulators, RNA processing factors, mTORC1 signaling components, and macrophage inflammatory genes that extend the known germline architecture of CH. We further anchor this phenotype within hematopoietic regulatory landscapes by demonstrating that associated variants are enriched in open chromatin regions across CD34⁺ HSPCs and aged bone-marrow HSPC multiome profiles, and by mapping prioritized genes to specific HSPC subpopulations and lineage programs.

Here, we used WGS data from 791,067 participants (including 481,079 UKB participants^19^ and 309,988 participants AoU ^20^ participants) to detect positive selection in aging blood with passenger mutations. By integrating results from a Genome Wide Association Study (GWAS), a gene-based Rare-Variant Association Study (RVAS), and sex-stratified and ancestry-stratified analyses, we mapped the common and rare germline architecture of passenger mutation burden and identified both shared loci and sex and ancestry specific signals. Complementing the common-variant architecture, rare-variant burden testing using deleterious missense masks highlighted signals at *MBD2* and related epigenetic and transcriptional regulators, implicating disrupted epigenetic reprogramming in hematopoietic stem and progenitor cell clonal fitness. In AoU, germline polygenic risk and observed passenger burden showed distinct phenotypic associations. Observed burden was more strongly associated with incident hematologic malignancies, all-cause mortality, and several nonhematologic outcomes than germline polygenic risk.

Spatial analyses of passenger mutation burden across U.S. regions revealed persistent geographic heterogeneity not explained by income, air quality, or chemotherapy exposure, pointing to additional unmeasured environmental or regional factors.

Overall, through parallel analyses of two population-scale biobanks, we use a quantitative measure of passenger mutation burden to characterize the genetic architecture of CH and its clinical sequelae.

## Results

We constructed a quantitative, driver-agnostic phenotype for clonal expansion by quantifying the burden of somatic passenger mutations in peripheral blood from whole-genome sequencing (WGS) of 791,067 samples across several ancestral backgrounds.

Somatic variants were identified and filtered using allele-depth (AD/DP) thresholds and germline exclusion criteria to minimize technical artifacts (Methods). Following recent analyses of passenger mutation burden ^14,16,18^, we then limited analysis to singletons as a conservative filter to deplete the calls of driver mutations, and we refer to these mutations as “passengers.”

### The common variant germline determinants of passenger mutation burden

We performed a GWAS in 481,079 UK Biobank participants and identified 45 genome-wide significant loci associated with passenger mutation burden (P < 5 × 10⁻⁸; Supplementary Figure S1, Supplementary Table S1). In a parallel GWAS in with 309,988 participants from the All of Us (AoU) Research Program we identified 16 genome-wide significant loci (Supplementary Figure S2).

Previous reports on the genetic epidemiology of CH have largely focused on European ancestry cohorts. To increase statistical power and the genetic diversity of the included participants we performed a meta-analysis combining UKB and AoU (791,067 total individuals) across 26,399,928 variants, identifying 81 genome-wide significant loci (**Figure 1A**; Supplementary Table S2). Of these, 39 replicated known CH loci and 42 were novel. Genomic control lambda was 0.981, indicating no test-statistic inflation. Among 1,078 genome-wide significant variants present and harmonizable in both cohorts, AoU and UKB effect estimates were highly correlated (Pearson r = 0.95) with concordant effect direction throughout (Supplementary Figure S3B).

**Figure 1.**
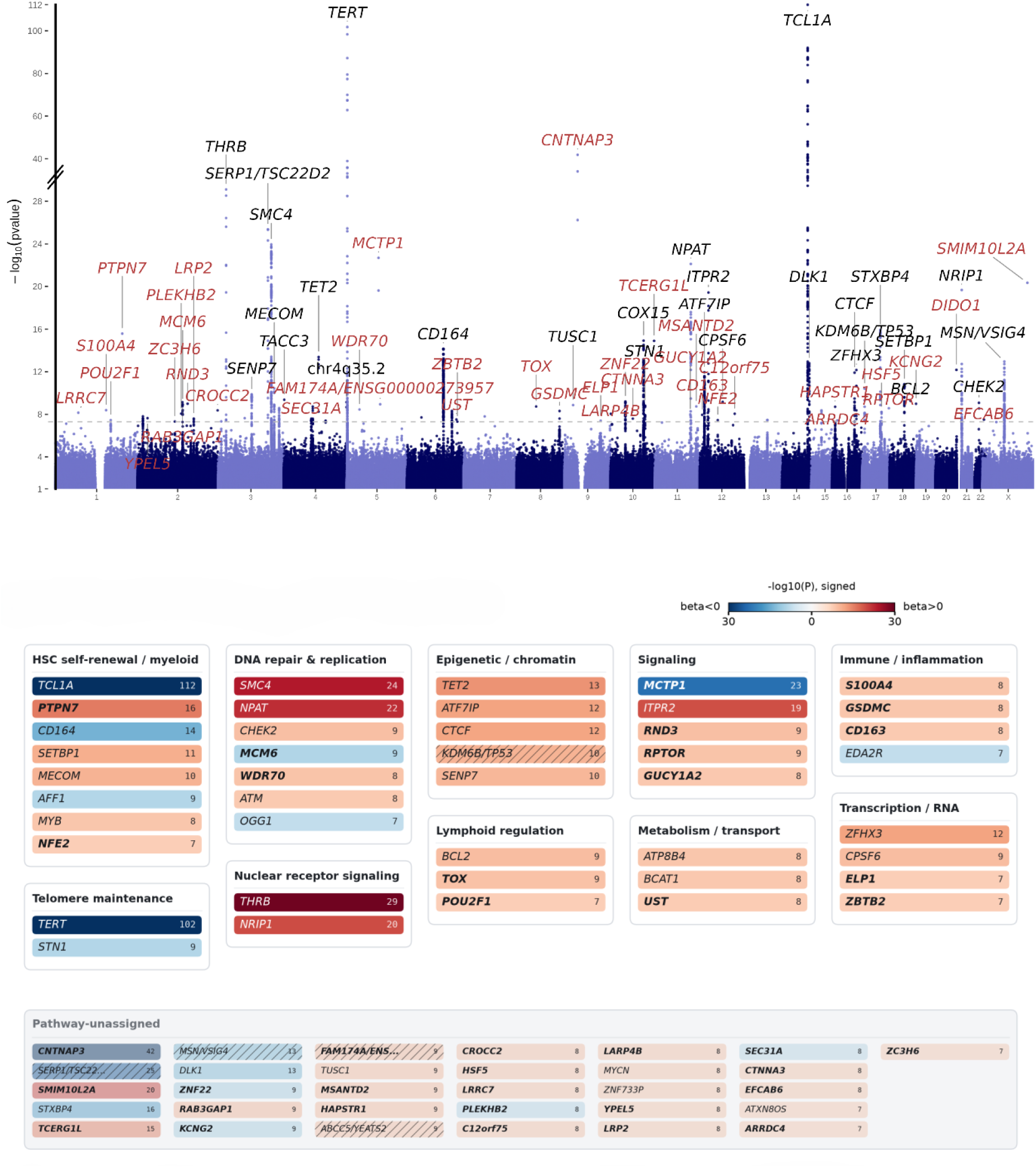
Genome-wide architecture of passenger mutation burden. **A**. Meta-analysis of passenger burden GWAS across 791,067 UK Biobank and All of Us participants, highlighting 81 genome-wide significant loci (P < 5 × 10⁻⁸). GWAS was performed with REGENIE in both cohorts, which were meta-analyzed with fixed-effects meta-analysis. Novel loci are highlighted in red text. **B.** Genome-wide significant loci from panel A (P < 5×10⁻⁸), annotated with nearest or empirical-Bayes (EB)-prioritized gene labels (Methods). The number in each box is the signed −log₁₀(P) for that locus (color scale: blue = negative β, red = positive β). Gene names in **bold** indicate novel genes; non-bold names indicate nearest-gene labels retained where EB prioritization was not resolved. Hatched boxes indicate loci with ambiguous, slash-delimited EB labels. Genes were grouped into pathways based on pre-existing CH/HSPC pathway-membership annotations used as a biology prior in the EB model; loci whose prioritized gene lacked prior pathway evidence are shown in the “pathway-unassigned” category.

To nominate causal genes at associated loci, we applied an empirical Bayes (EB) framework synthesizing evidence across coding sequence pathogenicity ^21^, Open Targets enhancer annotations ^22,23^, CD34⁺ chromatin accessibility ^24^, and eQTL colocalization ^25^ (Methods). This approach assigned high-confidence gene labels (posterior ≥ 0.9) to 60 loci, resolving several nearest-gene misattributions.

The meta-analysis recapitulated the canonical peaks at *TCL1A* (rs2887399, β = −0.040, P = 5.45 × 10⁻¹¹³) and *TER*T (rs2736100, β = −0.031, P = 1.61 × 10^-102^), concordant with recent reports on the germline genetic architecture of CH ^9,11,15,26,27^. The 39 replicated loci collectively span canonical CH biology including telomere maintenance (*TERT*), DNA damage response (*ATM*, *CHEK2*), apoptosis (*BCL2*), cell-cycle (SMC4), and hematopoietic stem and progenitor cell (HSPC) self-renewal (*TCL1A*, *MECOM*, *MYB*, *TET2*).

Among the 42 novel loci, several implicate biological mechanisms not captured by CH driver mutation-ascertained phenotypes. We highlight four novel loci with functional genomic lines of support.

At the *NFE2* locus, a common variant (rs35979828, MAF = 5.7%) was associated with increased passenger mutation burden (β = +0.016, P = 3.5 × 10⁻⁸). rs35979828 resides in a distal enhancer linked to *NFE2* ^28^and is associated with reduced NFE2 expression (β = −0.73, P = 1.8 x 10^-37^) in monocytes^29^, implicating reduced expression of *NFE2* in increased passenger burden. *NFE2* encodes a master transcription factor of erythroid and megakaryocytic maturation^30^. Mutated *NFE2* is recurrent in myeloproliferative neoplasms, and is linked to elevated risk of transformation to AML^31^. Thus, through common variant analysis, we discovered that variants that affect gene regulation recapitulate genes that have previously been discovered through recurrent coding mutations in diseased settings.

At the *MSANTD2* locus, a common variant (rs183501631; MAF = 6.2%, β = +0.023, P = 1.8 × 10⁻⁹) was associated with increased passenger burden. The lead variant overlaps a proximal enhancer cCRE and is predicted by ENCODE rE2G ^23^ to regulate *ESAM* (Endothelial Cell-Selective Adhesion Molecule) in CD34⁺ hematopoietic progenitor cells, suggesting that rs183501631 may modulate HSC activation and cycling through cis-regulatory control of ESAM, which is used as an HSC marker ^32^.

At the *PTPN7* locus the lead variant ((rs1413023105, MAF = 0.043%, β = +0.475, P = 2.6 × 10⁻^16^) is a rare, non-coding SNP ∼37 bp from a proximal-enhancer-like cCRE (EH38E2858479). *PTPN7* encodes a hematopoietic-restricted protein tyrosine phosphatase that regulates ERK/MAPK signalling in lymphocytes, and elevated *PTPN7* expression has been associated with poor overall survival in acute myeloid leukemia ^33^.

At the *CD163* locus (rs59730643, MAF = 0.34%, β = +0.090, P = 1.1 × 10⁻⁸), the EB framework reassigned the signal from the nearest gene *APOBEC1* to *CD163* based on cCRE linkage in myeloid progenitors. CD163 is a macrophage-restricted scavenger receptor for haptoglobin-hemoglobin complexes^34^ and a canonical marker of anti-inflammatory (M2-like) macrophage polarization^35^.

Additional novel loci implicate mTORC1 signaling (*RPTOR*), RNA processing (*CPSF6*, *TCERG1L*), and broader transcriptional regulatory programs. Taken together, meta-analysis of common variant analyses expands characterization of the inherited genetic architecture of CH, replicating established CH biology and nominating novel mechanisms for experimental follow-up (**Figure 1B**).

### Sex-stratified analysis reveals both shared and sex-specific germline determinants of somatic passenger burden

Stratifying our GWAS by sex, we identified 14 novel sex-specific loci that were not discovered in the non-stratified UKB GWAS. Female-specific loci included genes such as *RMND5B* and *ACTN1*, while male-specific loci included genes such as *OLIG3* and *ADCY2* (Supplementary Figure S4–S5, Supplementary Table S3–S4).

To characterize sex-specific and sex-shared germline determinants, we compared effect sizes at genome-wide significant loci between males and females, plotting βfemale against βmale with 95% confidence intervals (**Figure 2**). Overall, effect sizes were highly concordant across sexes (correlation = 0.89; Figure 2), with canonical loci including *TERT*, *TCL1A*, *DLK1*, and *DACT2* acting as sex-independent germline determinants of somatic passenger burden. The largest sex-specific signal in the dataset was *RMND5B* (rs192756388; β female = 0.189, β male = −0.011), which was absent from the non-stratified GWAS, indicating a signal entirely driven by females. *RMND5B* encodes a core subunit of the CTLH E3 ubiquitin-protein ligase complex ^36,37^. One locus showed stronger effects in males than in females: *OLIG3* (rs72975343; β female = 0.098, β male = 0.143), sitting above the diagonal in Figure 2. Several additional loci showed female-enriched effects, including *SMC4* (rs116205775; β female = 0.109, β male = 0.063).

**Figure 2.**
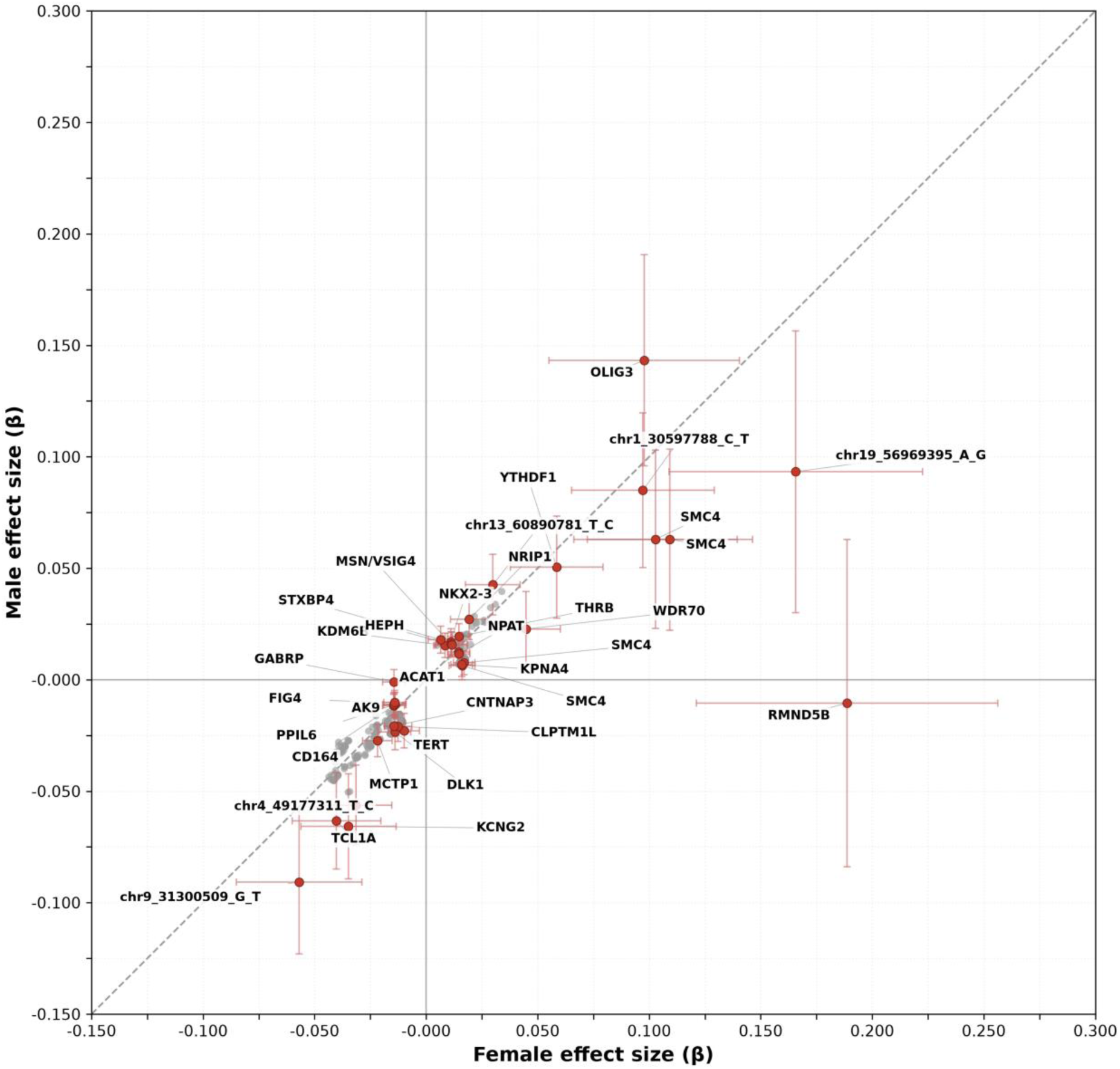
Sex-stratified germline effects on somatic passenger burden. Scatter of effect sizes at genome-wide significant SNPs comparing male and female GWAS in UK Biobank participants. Loci that appear off the diagonal and were not genome-wide significant in the non-stratified GWAS.

### Multi-ancestry GWAS identifies shared and ancestry-specific loci for somatic passenger burden

We performed ancestry-stratified GWAS in the All of Us Research Program cohort across six ancestry groups to characterize the genetic architecture of somatic passenger burden across diverse ancestries. This included the European (EUR; N = 178,805), African (AFR; N = 65,970), East Asian (EAS; N = 7,190), Admixed American (AMR; N = 61,533), Middle Eastern (MID; N = 1,194), South Asian (SAS; N = 3,968) and unassigned (not considered N = 35,772) cohorts.

We identified 194 genome-wide significant SNPs (p < 5×10⁻⁸) in total across all the ancestry cohorts. The large majority came from the EUR ancestry cohort (184 SNPs), with additional signals in AFR (8 SNPs) and EAS (2 SNPs). No genome-wide significant associations were observed in AMR, MID, or SAS, consistent with reduced statistical power due to sample size in those groups rather than the absence of genetic effects (**Supplementary Figure S6 - S7**). These 194 SNPs collapsed into 12 independent lead loci using a window-based approach (500kb), thus retaining the most significant SNP per region. The larger number of discovered loci in the EUR GWAS is due to two effects: 1) sample size, and 2) differences in the age at recruitment distribution across the ancestry strata (mean age = 52.5 years in EUR, versus 42.1– 50.1 years across the other ancestry groups; Supplementary Table S5). *TERT* (rs2736100) was the most significant locus genome-wide (β = −0.041, P = 1.0×10⁻³⁶) and *TCL1A* (rs2887399) the second strongest (β = −0.048, P = 3.7×10⁻³³), both showing negative effects in most ancestries (N = 6 for TCL1A and N = 5 for TERT, (**Figure 3A, Supplementary Table S6-S8**). Notably, while *TERT* showed attenuated effects in AFR and EAS that did not reach genome-wide significance — likely reflecting reduced power in those strata, *TCL1A* achieved nominal significance in both AFR (β = −0.025, P = 1.6×10⁻⁶) and AMR (β = −0.019, P = 0.005), supporting a stronger cross-ancestry germline effect at this locus.

**Figure 3.**
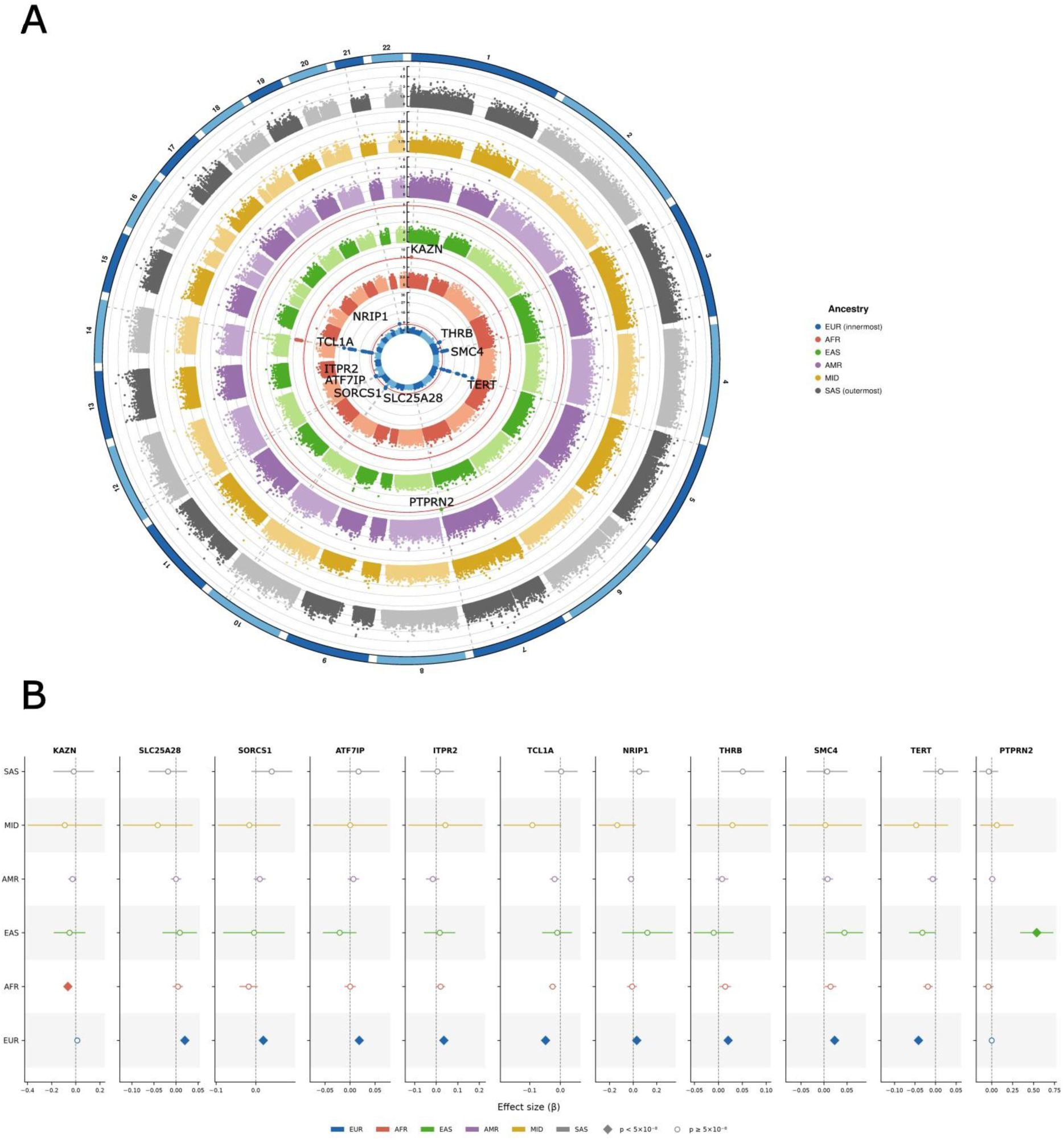
Multi-ancestry GWAS result of somatic passenger burden in All of Us cohort. **A**. Ancestry-stratified association results for mutation burden across six ancestry groups in All of Us (EUR, AFR, EAS, AMR, MID, SAS), comprising 309,988 participants with 194 genome-wide significant SNPs (P < 5 × 10⁻⁸). **B**. Significant SNPs collapsed to 12 independent lead loci. The forest plot depicts ancestry-specific effect sizes at the 12 key loci (e.g. TERT, TCL1A, TRIM59, ITPR2, KAZN, PTPRN2), highlighting shared signals with attenuated effects in non-EUR groups along with ancestry-specific associations.

Eight additional loci reached genome-wide significance in EUR, with effect sizes ranging from β = 0.018 (*ATF7IP*) to β = 0.036 (*ITPR2*) and consistent directions across ancestries (Figure 3B). Among these, *TRIM59* showed the strongest cross-ancestry support, with concordant positive effects in both AFR (β = 0.014) and EAS (β = 0.045), and *ITPR2* showed a positive effect in AFR (β = 0.020). The remaining loci - *SLC25A28*, *SORCS1*, *NRIP1*, *THRB*, *KPNA4*, and *ATF7IP*, were EUR-predominant, with consistent but attenuated effects in other ancestry groups that did not reach significance, likely reflecting limited power outside EUR.

The only AFR-specific genome-wide significant locus was *KAZN*/*TMEM51* ( rs4661322; β = −0.064, P = 2.8×10⁻⁸). It is an intergenic common variant between *KAZN* and *TMEM51* (AF ≈0.92). The locus sits in a CEBPA-bound regulatory element and a distal enhancer, though gene prioritization remained challenging. In EUR, the same SNP showed an effect in the opposite direction (β = +0.013, P = 0.078). The most striking ancestry-specific locus was *PTPRN2* (rs73171533), which reached genome-wide significance exclusively in EAS (β= 0.538, P = 3.5×10⁻⁸) with a large effect size and low alternate allele frequency in EAS (AF = 0.0065).

### Rare variant burden testing identifies germline coding associations with clonal haematopoiesis

To identify rare coding variant associations with somatic passenger burden, we performed gene level rare variant burden testing using REGENIE (Methods) on whole genome sequencing data from the UK

Biobank participants. Variants were aggregated by gene and tested using burden masks defined by AlphaMissense-predicted ^21^ deleterious missense variants at two score Genes reaching the exome-wide significance threshold of p < 2.5×10⁻⁶ were considered significant (Methods).

We observed that 37 genes reached exome-wide significance (Supplementary Table S10). Most were known CH drivers (e.g., *DNMT3A, SRSF2, TET2, SF3B1, GNB1, TP53, IDH2, PRPF8, GNAS, KRAS, CBL, SRSF1, MTA2*) or immunoglobulin-region hits, providing internal validation that the passenger burden phenotype captures clonal selection biology. After exclusion of somatic CH drivers and IG-region signals, 11 genes remained as candidate rare germline determinants of clonal expansion (**Figure 4-B**, Supplementary Table S9). We observed that deleterious variation at *MBD2* (log₁₀p = 18.51, β = 0.353, SE = 0.056) was associated with increased mutation burden. *MBD2* encodes a methyl-CpG binding domain protein that recruits the NuRD (nucleosome remodeling and deacetylase) chromatin remodelling complex to mediate epigenetic silencing at methylated promoters (**Figure 4C**). *DNMT3A* and *TET2,* the two most frequently mutated CH driver genes, write and erase methylation, whereas *MBD2* reads and interprets methylation marks and couples them to chromatin regulation. Thus, germline and somatic variation converge on disruption of methylation-dependent regulation, spanning the writing, erasing, and reading of DNA methylation marks. *PRKACB* (log₁₀p = 13.97, β = 0.652, SE = 0.082) encodes a catalytic subunit of protein kinase A, the effector immediately downstream of cAMP in the G-protein → adenylyl cyclase → cAMP → PKA cascade. *PUF60* (log₁₀p = 11.21, β = 0.566, SE = 0.093) is a poly-U-binding factor that acts in the same 3′ splice-site machinery as the somatic splicing drivers *SF3B1, SRSF2, and U2AF1* (**Figure 4D**). Overall, *MBD2* and *PUF60* extend canonical somatic CH pathways of methylation regulation and RNA splicing to inherited determinants, while *PRKACB* implicates cAMP/PKA signalling and connects rare germline variation to the common-variant mTORC1 signal at *RPTOR*.

**Figure 4:**
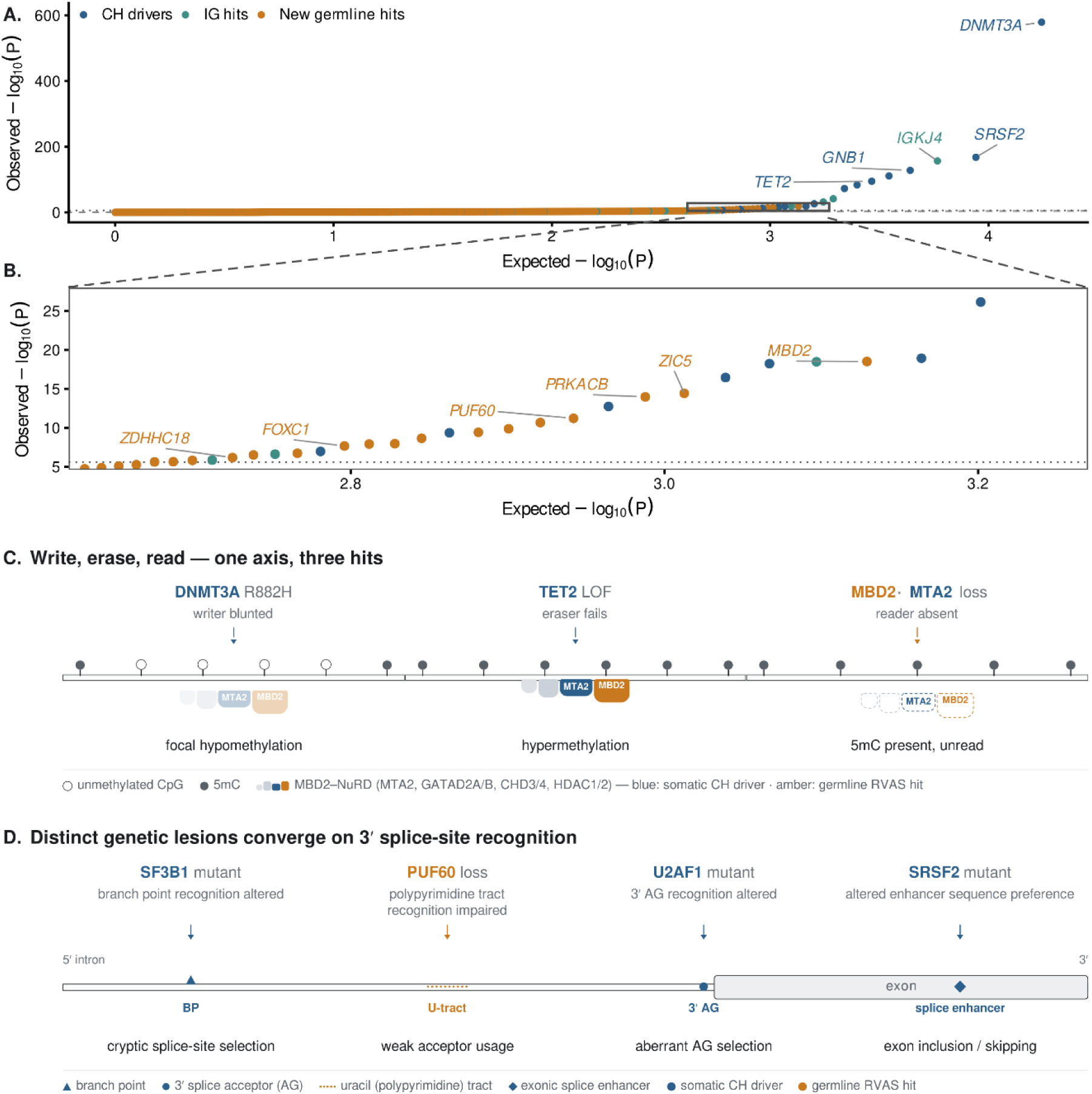
Gene-based rare-variant association QQ plot for somatic passenger burden. **A**. Expected and observed −log₁₀(P) values for gene-level tests, with genes coloured as somatic clonal hematopoiesis (CH) drivers, immunoglobulin (IG) hits, or new germline hits. The dotted horizontal line marks the exome-wide significance threshold (P = 2.5 × 10⁻⁶) **B.** Magnified region highlighting the upper tail of the new germline-hit category and selected IG-region signals. **C. M**ethylation-regulation axis at a representative CpG-rich locus. Somatic DNMT3A R882H blunts methylation writing, producing focal hypomethylation; somatic TET2 loss-of-function impairs methylation erasing, producing hypermethylation; germline MBD2 deleterious variation (acting through the MBD2–NuRD complex: MTA2, GATAD2A/B, CHD3/4, HDAC1/2) impairs methylation reading, leaving 5-methylcytosine marks present but uninterpreted. **D.** 3′ splice-site recognition axis, SF3B1 alters branch-point recognition; germline PUF60 impairs polypyrimidine-tract recognition; U2AF1 alters 3′ AG recognition; SRSF2 alters exonic splice-enhancer preference.

### Epigenomic Context of Regulatory GWAS loci

To contextualize the regulatory landscape of the prioritized germline CH loci, we assessed the enrichment of fine-mapped SNPs within open chromatin regions of HSPCs (**Figure 5A**). Using both 5 kb proximity and access-weighted overlap metrics, compared to a matched null, prioritized SNPs were highly enriched for regions accessible in young HSPC, aged HSPC, and CD34+ HSPC aggregate chromatin profiles ^38^, as well as in stimulated HSC states (PBS, TNF, and LPS). The access-weighted overlap metric yielded higher enrichment estimates (with > 2.0–3.0 fold change over null) compared to proximity-based overlap, suggesting that the association signal is concentrated within functionally active regulatory elements rather than co-localizing with open chromatin by genomic proximity alone.

**Figure 5:**
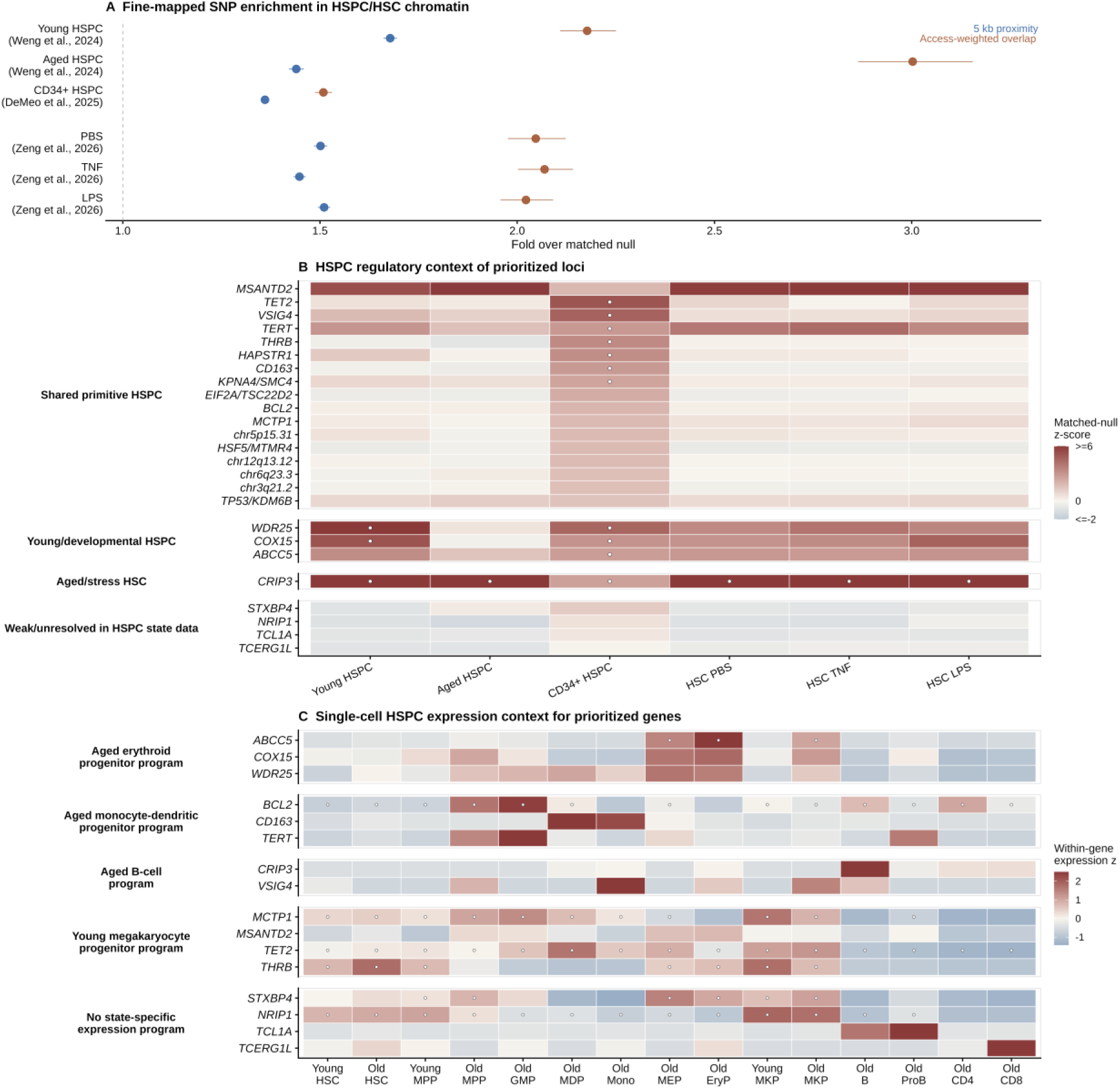
Epigenomic and HSPC cell-state context of prioritized CH loci. **A.** Enrichment of fine-mapped GWAS SNPs within the open chromatin regions of HSPCs across multiple reference datasets (young HSPC, aged HSPC, CD34⁺ aggregates, and stimulated HSC states), comparing 5 kb proximity-weighted accessibility (blue) to access-weighted overlap (orange) metrics; horizontal bars represent the resulting 95% confidence interval. **B.** Locus-level chromatin accessibility z-scores (capped at −2 to 6) across six HSPC reference states, classifying loci into shared primitive HSPC, young/developmental, and aged/stress groups. Open circles mark resource-level significance (q ≤ 0.10). **C**. Gene-level expression z-scores across 15 immunophenotypically defined HSPC subpopulations^39^, assigning loci to specific lineage types. Open circles mark genes expressed in ≥25% of cells within that subpopulation.

We then characterized the HSPC regulatory context of each prioritized locus using locus-level chromatin accessibility across the six HSPC reference conditions (**Figure 5B**). Most (n = 6) of the prioritized loci exhibited chromatin signal across multiple HSPC states and were classified as shared primitive HSPC loci, indicating that the regulatory elements harboring these variants are broadly active in primitive hematopoiesis. Three loci showed state-specific regulatory activity (*WDR25, COX15,* and *ABCC5*) and were preferentially accessible in young HSPC chromatin (Young/developmental HSPC group). In contrast, *CRIP3* showed selective accessibility in aged and stress-conditioned HSCs (Aged/stress HSC group). This age-stratified chromatin patterning suggests that aging remodels the accessible regulatory landscape of HSPCs, rendering distinct sets of germline variants functionally active or dormant at different life stages.

To resolve the cell-state specificity of prioritized CH loci genes, we examined gene expression across 15 immunophenotypically defined HSPC subpopulations (**Figure 5C**). Across these states, genes with young/developmental chromatin profiles tended to show highest expression in old erythroid progenitors, whereas loci such as *BCL2, CD163,* and *TERT* were enriched in old monocyte dendritic progenitors and monocytes, implicating myeloid-biased progenitor states as a major transcriptional context. *CRIP3* and *VSIG4* mapped to distinct lymphoid and myeloid programs, and several loci with broad primitive HSPC chromatin accessibility (including *TET2, MSANTD2, THRB*, and *MCTP1*) were assigned to a young megakaryocyte progenitor program. Overall, through integration with HSPC chromatin and expression profiling, we observed substantial heterogeneity in the regulation and expression of CH germline risk genes.

### Phenome Wide Association Study of Germline CH burden and Somatic Passenger Burden in the All of Us Research Program

We then asked whether germline risk for passenger mutations also conferred risk for common aging-related diseases. We performed a phenome-wide association study (PheWAS) in European ancestry (EUR) participants in the All of Us Research Program of 30 incident, common aging-related diseases in order to assess the phenotypic consequences of both inherited clonal haematopoiesis (CH) liability and measured passenger burden. We included both a cross-sectional case-control design and an incident disease analysis. To mitigate over-fitting, we trained a polygenic risk score of CH in UKB and then used this score to generate inherited risk predictions in All of Us.

The largest risk effect we observed was between germline risk and incident AML (β = 0.04), although due to a small number of cases the standard errors are large. We observed significant associations between the PRS and non-hematologic phenotypes, including obesity (OR 1.025, 45,191 cases, P=4.8×10⁻⁵; HR 1.013, P=9.8×10⁻³) and a protective effect for asthma (OR 0.975, 27,074 cases, P=3.3×10⁻⁴; HR 0.967, P=3.7×10⁻⁷). Protective associations were also observed with melanoma of skin (OR 0.943, 3,135 cases, P=1.8×10⁻³), dementias (OR 0.932, 2,160 cases, P=1.7×10⁻³), Alzheimer’s disease (OR 0.904, 633 cases, P=1.2×10⁻²), in concordance with a recent report describing a protective effect of CHIP on AD risk ^40^. Concordant with previous analyses of barcode-CH ^15^, we found little enrichment of CAD phenotypes (Figure 6, Supplementary Table S11).

**Figure 6:**
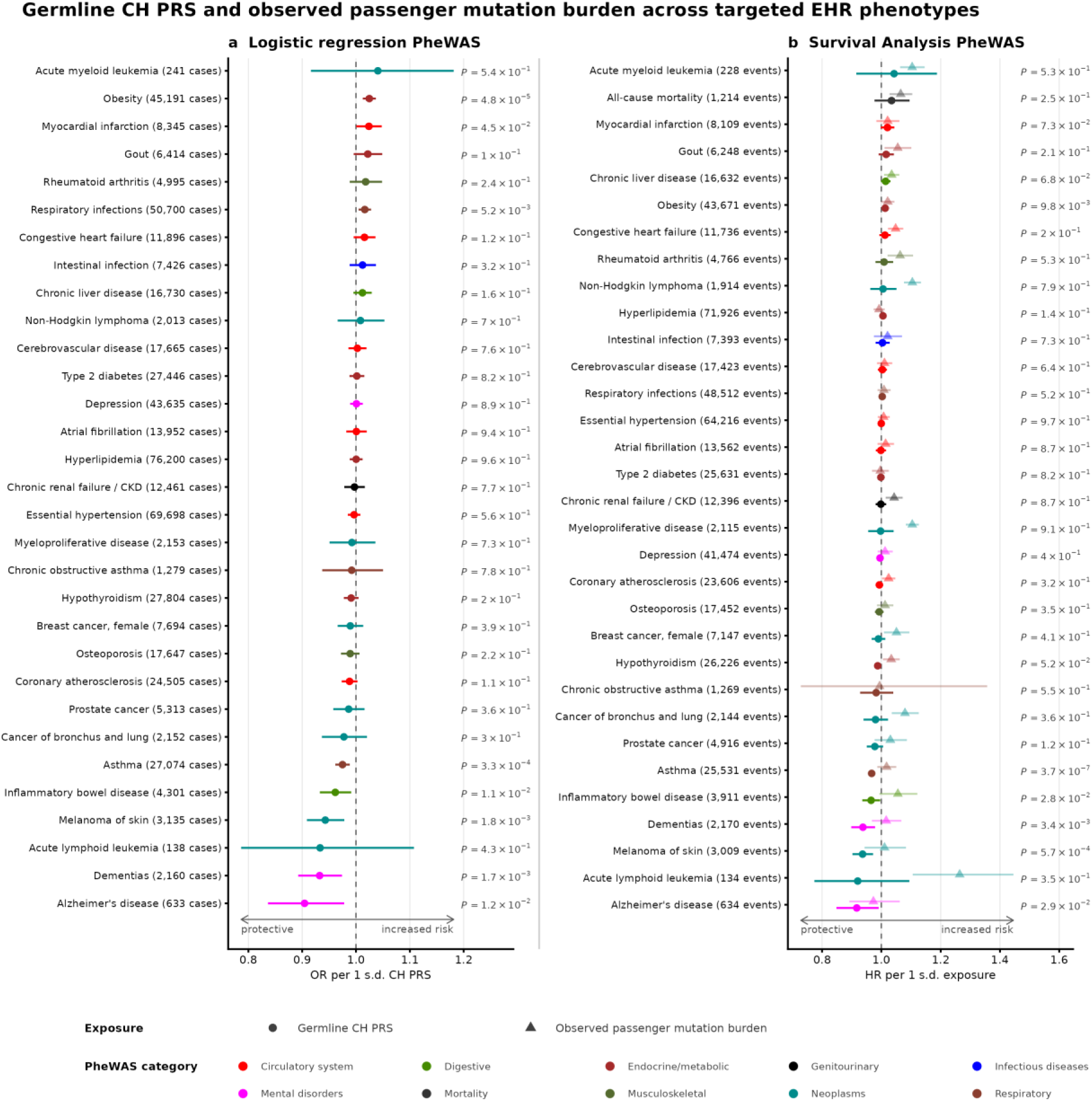
Phenome-wide associations of germline CH risk and somatic passenger burden in All of Us. Phenome-wide association results for two aspects in EUR All of Us participants - a polygenic risk score (PRS) for clonal hematopoiesis derived in UKB, and observed somatic passenger burden at blood draw. Odds ratios (OR) and hazard ratios (HR) for case control and incident disease are plotted with point estimates and 95% confidence intervals.

Larger effect sizes between observed passenger mutation burden and incident hematologic malignancy were observed than with the germline risk instrument. Increased risk was observed across hematologic outcomes: MPN HR 1.10 (586 events, P=3.2×10⁻²²), non-Hodgkin lymphoma HR 1.10 (418 events, P=8.3×10⁻¹⁴), AML HR 1.10 (93 events, P=2.7×10⁻⁷), and all-cause mortality HR 1.07 (1,179 deaths, P=6.4×10⁻⁴). Beyond haematologic outcomes, passenger burden was also associated with post-draw incident lung cancer (HR 1.08, 725 events, P=3.9×10⁻⁴), chronic kidney disease (HR 1.04, 3,481 events, P=2.7×10⁻³), chronic liver disease (HR 1.03, 5,269 events, P=7.3×10⁻³), and breast cancer (HR 1.05, 1,176 events, P=0.017) (**Figure 6**, Supplementary Table S12).

The divergence between the germline PRS and observed passenger mutation burden underscores the presence of environmental effects that cause variation in both disease and passenger mutation burden. A subset of phenotypes displayed antagonistic genetic and environmental effects – germline risk for CH protects against lymphoid malignancy, but mutation burden is strongly enriched in carriers of lymphoid malignancy. The opposing associations are consistent with germline CH liability buffering ALL risk associated with acquired passenger burden. Overall, the divergence between inherited risk and observed burden represents an opportunity to better characterize the environmental exposures to CH.

### Geographical Heterogeneity of Mutation Burden

After characterizing the genetic determinants of CH, we then studied the environmental contributions to passenger burden. To characterize the contribution of environmental and geographic factors to passenger burden, we performed a spatial regression analysis across geographic regions of the United States in the AoU cohort. After residualizing passenger burden for age, sex, smoking status, genetic ancestry principal components and technical sequencing covariates, we linked the individual-level phenotype to geographic location of the participants using three-digit ZIP code (ZIP3) regions. The final participant-level cohort for geographic analysis comprised 330,056 individuals distributed across 804 distinct ZIP3 regions; after excluding ZIP3 areas with fewer than 10 analytical participants, 471 ZIP3 regions were retained for area-level analyses (Figure 7A).

**Figure 7:**
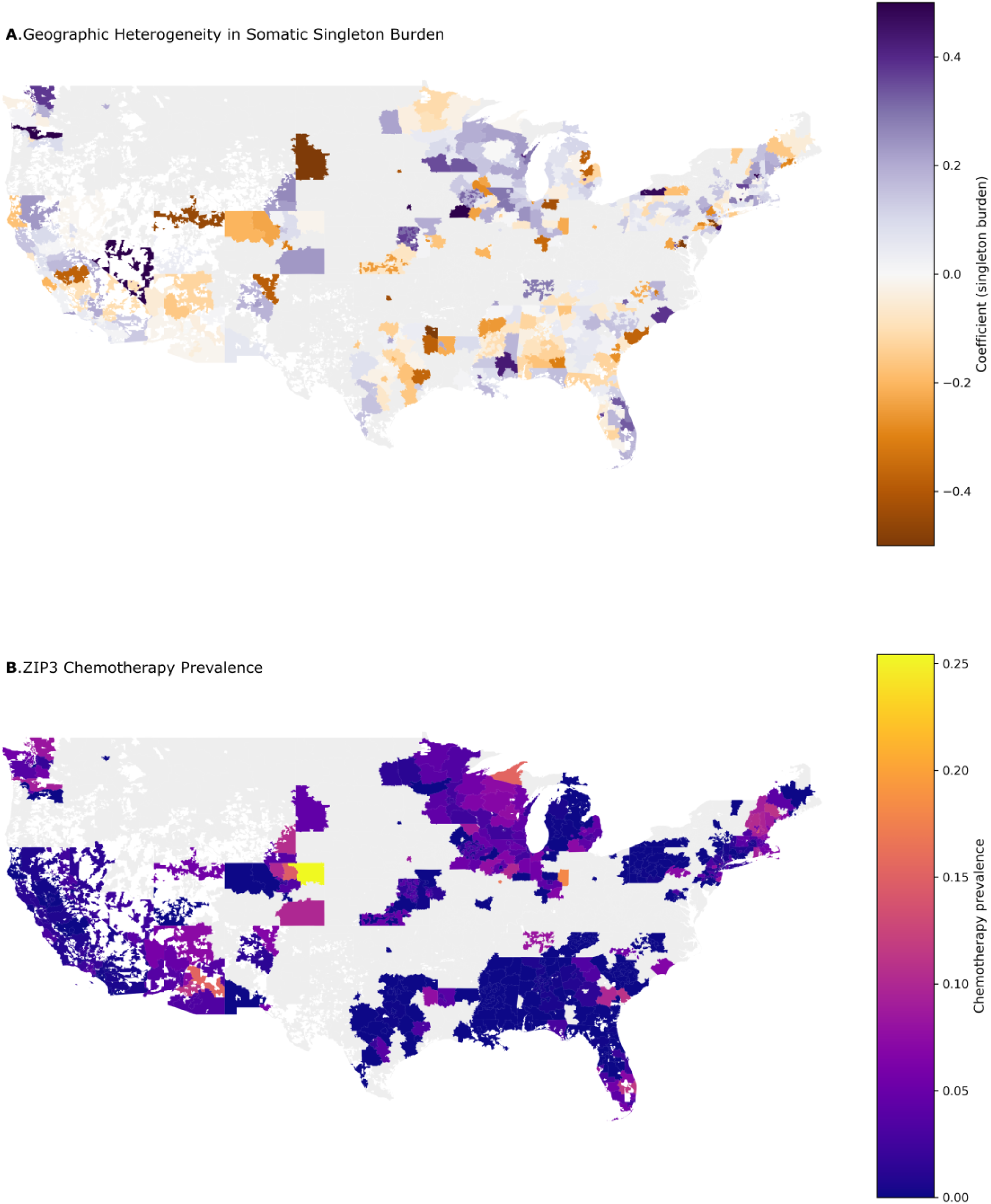
Geographic heterogeneity of passenger mutation burden in All of Us. **A**. Spatial patterns of residualized passenger mutation burden across three-digit ZIP code (ZIP3) regions in the United States of America, after adjustment for ancestry principal components and sequencing covariates. The first map shows mean residual burden across 471 ZIP3 regions having at least 10 participants. **B**. Spatial autoregressive model results (spatial lag and spatial error). This includes estimates of spatial parameters (ρ, λ) and the effect of chemotherapy prevalence, showing an improved fit over non-spatial OLS and significant residual spatial autocorrelation.

We tested whether ZIP3-level socioeconomic context (median household income, ACS 2022), air quality (Air Quality Index, EPA), and chemotherapy exposure prevalence explained the observed geographic heterogeneity. In non-spatial ordinary least squares (OLS) models we saw that none of the three candidate area-level covariates explained significant residual variance in the geographic pattern; income and AQI explained little residual variance (details here) (Supplementary Figure S8). The largest effect size was observed for chemotherapy prevalence (Figure 7B), though the effect was difficult to distinguish from a null model due to limited statistical power.

Residual Moran’s I remained positive after adjustment for all three covariates (I = 0.188, P = 0.004), indicating that the spatial clustering of passenger mutation burden persisted beyond these external variables. We therefore fit formal spatial autoregressive models within the largest connected ZIP3 contiguity component (119 ZIP3 areas representing 60,674 participants). Both a spatial lag model and a spatial error model substantially improved fit over OLS (AIC: OLS = −85.9; spatial lag = −92.6; spatial error = −95.2), with the spatial error model performing best. The spatial error parameter was strongly significant (λ = 0.386, P = 1.5 × 10⁻⁴), and the spatial lag parameter was similarly robust (ρ = 0.351, P = 7.0 × 10⁻⁴). Chemotherapy prevalence had the largest effect size in both spatial models (spatial error β = 0.030, P = 0.056) (Supplementary Table S13). Overall, these results indicate that the geographic heterogeneity of passenger mutation burden is incompletely explained by income, air quality, or chemotherapy exposure in participants, and instead reflects substantial spatially structured variation likely driven by additional unmeasured environmental factors.

## Discussion

Humans develop somatic mosaicism as they age, yet how and why remains incompletely understood. In this work, we used whole-genome sequencing data from 791,067 individuals across the UK Biobank and All of Us Research Program to construct a quantitative, driver-agnostic CH phenotype. In a UKB–AoU multi-ancestry meta-analysis, we identified 81 genome-wide significant common-variant loci, of which 42 had been previously unreported. The genetic architecture of the overall meta-analysis masked substantial heterogeneity in the genetic architecture: GWAS stratified by sex and ancestry yielded loci not detected in the primary analysis (14 sex-specific loci, 4 ancestry-specific loci). Permitted by the parallel analysis of two population-scale biobanks, we mapped the disease correlates of both polygenic risk and the observed mutation phenotype.

Our results yield study design considerations for genetic epidemiology of CH. An important advancement from this study is that a driver-agnostic, continuous passenger burden phenotype yields a greater number of locus discoveries than driver-mutation case–control GWAS despite a smaller overall sample size than recent driver-based studies ^27^. We found that the quantitative passenger burden phenotype recapitulated canonical CH loci while also identifying 42 novel loci.

The expanded genetic discovery afforded by passenger mutation burden reveals striking convergence of common germline variants, rare coding variants, and somatic CH drivers on pathways that regulate HSC fitness in aging blood. We highlight four examples of this somatic-germline convergence. First, the canonical CH driver landscape is dominated in frequency by two epigenetic regulators, *DNMT3A* and *TET2*, which have key functions as a methylation writer and eraser respectively. Through our rare-variant analysis, we discovered deleterious variation in *MBD2,* which expands the methylation regulation axis through its role as a methylation interpreter. Similarly, we observed the convergence of regulators of repressive chromatin and histone modifications, including somatic *ASXL1,* with germline loci including *ATF7IP, SENP7,* and *KDM6B.* Third, we observed genome maintenance and DNA repair as a third convergent mechanism, including somatic mutations in *PPM1D*/*TP53*, along with germline regulation of *TERT, CHEK2, MCM6,* and *WDR70.* Finally, we observed core spliceosome machinery as a fourth axis, including somatic drivers in *SF3B1, SRSF2,* and *U2AF1*, along with deleterious germline variation in *PUF60*.

Our genetic discovery also nominates novel signalling pathways that have been previously uncharacterized in CH biology. Common variant analyses implicated regulation of *RPTOR,* a component of mTOR complex 1 ^41^, and *PTPN7,* a protein-tyrosine phosphatase that regulates the MAP kinase pathway ^42^. Rare-variant analyses identified deleterious coding variation in *PRKACB*, which encodes a subunit of protein kinase A. Remarkably, *PRKACB* has been shown to phosphorylate *RPTOR*, leading to decreased mTORC1 activity ^43^, implicating common and rare convergence in regulation of the mTORC1 pathway in HSCs.

Our results also reveal how germline architecture differs across sex and ancestry. While most genome-wide significant loci showed concordant effects between males and females, *RMND5B* exhibited a female-specific association and was absent in the non-stratified analysis. Ancestry-stratified analyses yielded an AFR-specific signal at *KAZN/TMEM51* and an EAS-specific, large-effect association at *PTPRN2* underscoring the increase in power afforded through population-scale multi-ancestry analysis.

The phenome-wide analyses reveal divergence between inherited CH liability and observed mutation burden. Germline CH PRS were associated with several non-hematologic outcomes, including increased obesity risk and reduced risk of asthma, melanoma, dementia, and Alzheimer’s disease, concordant with reports of protective associations between CHIP and Alzheimer’s disease risk ^40^. Observed passenger burden exhibited stronger associations with incident MPN, non-Hodgkin lymphoma, AML, all-cause mortality, and several non-hematologic outcomes (lung cancer, chronic kidney disease, chronic liver disease, breast cancer). These findings suggest that epidemiologic analyses relating CH to disease should consider inherited CH liability, which may confound associations between acquired clonal burden and disease risk. Furthermore, the contrast in phenotype associations between the CH PRS and observed passenger burden indicate that examining CH without consideration of germline risk is likely to mask heterogeneity in underlying risk for incident disease.

The contrast between germline PRS and passenger burden also has implications for AML risk prediction and CH risk stratification tools such as CHRS ^44^. Existing approaches rely on driver mutation status, clone size, and limited clinical covariates. Our findings indicate that incorporating germline CH PRS and continuous passenger burden measures could improve risk stratification, for example by distinguishing individuals with modest driver-detected clones but high passenger burden (and thus elevated near-term risk) from those with similar drivers but low overall burden.

Spatial analyses in the All of Us cohort revealed geographic clustering of passenger mutation burden that persisted after adjustment for area-level income, air quality, and chemotherapy exposure prevalence and was better captured by spatial autoregressive models than by non-spatial OLS. Thus, indicating that unmeasured environmental factors mediate substantial geographic heterogeneity in CH and motivate future epidemiologic work integrating richer exposure data (e.g., radiation, fine-scale pollution metrics, or healthcare access).

Several limitations should be considered when interpreting these results. First, statistical power for ancestry-stratified analyses was highly uneven; the absence of genome-wide significant loci in three ancestry cohorts most likely reflects limited sample size rather than a true absence of genetic effects, and the EUR-enriched loci that show only nominal associations in other ancestries should be interpreted in this light rather than as evidence of EUR-specificity. Second, differences in age-at-recruitment distributions across ancestry strata (with EUR participants oldest on average) may have contributed to differential power independent of sample size, complicating cross-ancestry comparisons of effect sizes and discovery rates. Third, the geographic analyses are correlational and cross-sectional; although spatial autoregressive modelling demonstrates residual clustering beyond the covariates tested, the specific unmeasured environmental exposures underlying this pattern remain unidentified. Finally, PheWAS and incident-disease analyses were conducted in All of Us EUR participants only; extending these phenotypic association analyses to non-EUR ancestry groups as sample sizes grow will be critical for assessing generalizability.

Overall, through population-scale analysis of passenger mutations, we advance characterization of the etiology of CH. We anticipate that our observations will improve CH surveillance and risk prediction for AML and other aging-related diseases at the population level.

## Methods

### Phenotype Definition

We used blood-derived WGS data from UKB and the AoU to construct quantitative measures of somatic mutation burden. Somatic single-nucleotide variants were called from blood WGS using a joint-calling pipeline with standard filters on depth and mapping quality, followed by additional read-level and population filters described below. To isolate true somatic singletons from germline variants and sequencing artefacts, we applied a multi-stage filtering strategy. First, we retained only single-nucleotide polymorphisms observed in exactly one genome across the cohort, which are termed as singletons.

Second, for each candidate singleton we required the alternate allele to be supported by 4–8 reads out of 30-fold total depth. This filtering criterion corresponds to a variant allele fraction of roughly 13–26 percent, which is consistent with mutations carried by clones present in about one in four to one in two blood cells. We then filtered for a total depth of 20–50.

To ensure that retained singletons represent true somatic events rather than extremely rare germline variation, we removed any variant that appeared in large germline reference of the TOPMed databases. The resulting set contained singletons with appropriate read-level support, local depth, and no evidence of germline variants. We then used a generalized linear model with a Poisson likelihood to compute the residual mutations relative to a baseline of age at recruitment, sex, and smoking, which was then used as the quantitative phenotype in UKB.

In AoU, we called passenger mutations on chr21 to reduce cost relative to whole-genome calls. Using the same somatic filtering criteria, we counted putative passenger mutations on chromosome 21, restricting to blood-derived samples. We excluded all mutations on the short-arm of the chromosome, due to the abundance of highly repetitive sequence. To construct a covariate-adjusted phenotype, we fit a generalized linear model with a Poisson likelihood. We included age at recruitment, sex, and ever-smoked status as covariates and computed residuals. These residuals were used as the quantitative trait in the AoU genome-wide association analysis.

### Common Variant GWAS in UK Biobank

For UKB, we applied a REGENIE two-step linear mixed-model workflow for quantitative traits to the GRCh38 merged QC-passed genotype panel^45^. The covariate set included three spline terms for age at recruitment, sex, ever-smoked status, ten genetic principal components derived from genome-wide genotypes, and individual-level measures of sequencing depth, coverage, and estimated contamination. Sex and ever-smoked were coded as categorical covariates. We enforced a minimum minor allele count threshold of six hundred and excluded variants falling in regions of poor mappability, or those that were present in contig differences between GRCh38 and GRCh37. We additionally excluded significant loci where the lead variant had a pvalue that was several (at least 5) order of magnitude smaller than the second smallest pvalue in the locus (500kb window), as we found these loci were enriched for artifacts.

### Common Variant GWAS in All of Us

In the All of Us cohort, we applied the same two-step REGENIE^45^ framework. Covariates for the AoU GWAS included three spline terms for age at recruitment, sex, ever-smoked status, ten genetic principal components derived from genome-wide genotypes, and individual-level measures of sequencing depth, coverage, and estimated contamination. We excluded variants based on the same criteria as in UKB.

### Meta-Analysis

We meta-analyzed the UK Biobank and All of Us GWAS using fixed-effect inverse-variance meta-analysis implemented in the gwasplot package ^46^. We removed variants in regions with known mapping difficulties (hg19diff, UCSC unusual regions, and Genome Reference Consortium exclusion regions). We then performed fixed-effect meta-analysis after harmonizing alleles between studies and computed pooled effect sizes and standard errors using inverse-variance weighting. Genome-wide significance was defined at P < 5 × 10⁻⁸.

### Causal Gene Prioritization

We performed SNP-to-gene prioritization by defining genome-wide significant lead loci from meta-analysis variants with P < 5 × 10⁻⁸ using 500 kb windows. For HSPC gene linking, we expanded candidate genes per locus such that it included the nearest and reference-overlap genes, optionally the blood rE2G-linked genes, and protein-coding cis genes within 250 kb. We then summarized HSPC expression and promoter accessibility along with the lead-variant and promoter peak overlaps, blood and CD34 rE2G evidence, and PIP-TWAS support. This was later combined into interpretable context and anchor scores that captured compatibility with HSPC biology.

The final effector labels were obtained from an empirical-Bayes model that combines an annotation prior with a genetic evidence term. The prior uses features that are independent of GWAS test statistics which includes the local context, expression/accessibility, rE2G and cCRE links, lead-variant consequence and enhancer annotations, and CH/HSPC pathway membership, while ACAT, colocalization, fine-mapping PIP^47^, and PIP-TWAS come into picture only through the genetic evidence layer. For pathway summaries, including the HSC pathway context figure, we group empirical-Bayes effector labels according to the existing CH/HSPC pathway-member flags, leaving loci without such prior evidence in a pathway-unassigned category.

### Sex-stratified GWAS of Passenger mutation burden

We split the UK Biobank cohort into female and male subsets based on the genotype defined sex and re-ran the two-step linear mixed-model GWAS workflow separately for each sex stratum, using the same QC-passed GRCh38 genotype panel and covariate framework as in the non-stratified analysis, except that sex was omitted as a covariate because stratification handled this dimension directly.

Within each sex-stratified GWAS, genome-wide significant variants were defined at P < 5 × 10⁻⁸. Lead loci and their associated summary statistics were obtained separately for the female-only and male-only GWAS using identical QC and windowing parameters.

### Ancestry stratified GWAS in All of Us

We performed ancestry-stratified GWAS in the All of Us Research Program to characterize germline associations with somatic passenger mutation burden across diverse genetic backgrounds. Analyses were restricted to 309,988 participants with blood-derived WGS, a non-missing residualized passenger burden phenotype, complete covariates, and ancestry assignments (of which around 35,000 individuals did not have ancestry information). The phenotype was the residualized mutation-burden measure described above in all ancestry strata.

Participants were assigned to six ancestry groups which included the European (EUR), African (AFR), East Asian (EAS), Admixed American (AMR), Middle Eastern (MID), and South Asian (SAS) by using the All of Us genetic ancestry pipeline based on principal-component clustering, and only individuals with high-confidence ancestry labels were retained for stratified analyses. Approximate stratum sizes were EUR = 178,805, AFR = 65,970, EAS = 7,190, AMR = 61,533, MID = 1,194, and SAS = 3,968. Within each ancestry group, we applied the same two-step linear mixed-model framework used for the primary All of Us GWAS. We applied identical variant-level QC within each stratum.

Genome-wide significance was defined at P < 5 × 10⁻⁸ within each ancestry-stratified GWAS. Across all six strata, genome-wide significant SNPs were pooled and collapsed into independent lead loci using the same 500 kb window-based procedure where variants were grouped by chromosome and position, and the most significant SNP within each 500 kb window was retained as the lead, with ancestry-specific effect estimates and allele frequencies recorded for that variant.

### Rare variant burden analysis (RVAS) in UK Biobank

To identify rare coding germline determinants of somatic passenger mutation burden, we performed gene-level rare-variant burden testing in UK Biobank WGS data using REGENIE^45^. The exposure phenotype was the same phenotype used for the common-variant GWAS in UK Biobank.

We annotated variants with AlphaMissense scores and constructed two nested missense burden masks per gene: a stringent mask including missense variants with AlphaMissense score ≥ 0.564 (likely pathogenic classification), and a more inclusive mask including missense variants with score ≥ 0.340 (capturing likely and ambiguous deleterious candidates), both restricted to alternate-allele frequency of < 0.001 in the analysis cohort. Burden masks were built using maximum-mask burden construction with alternate-allele frequency thresholds up to 0.5%, and REGENIE step-2 tests combined burden and dispersion components (SKAT-O and ACAT-family) to obtain gene-level omnibus P-values (GENE_P) for the phenotype.

Lead genes were defined as those with GENE_P P < 2.5 × 10⁻6 (LOG10P ≥ 5.602) in the RVAS results. For each significant gene, we identified the mask contributing the strongest association from the REGENIE strongest-mask field and used the corresponding per-gene variant sets from the local mask-definition and annotation tables, mapping them to the appropriate AlphaMissense score thresholds (≥ 0.34 and ≥ 0.564). This procedure yielded a set of “strongest-mask” variants for each exome-wide significant gene, which were used for downstream burden-effect and dose-response summaries.

For each exome-wide significant gene, we extracted the additive burden effect size and direction from the REGENIE additive tests, as well as positive-versus negative-direction burden statistics from the signed burden components. To assess the AlphaMissense dose-response, we compared per-allele burden effects for each gene under the broader (score ≥ 0.34) versus stricter (score ≥ 0.564) missense mask, and used this pattern together with (i) prior knowledge of somatic CH-driver status and (ii) locus context (regional clustering of signals, location in the MHC or known segmental duplications, and whether significance was driven by burden or dispersion) to tier genes into high-confidence inherited candidates, ambiguous signals, and likely artifacts. Genes that were likely artifacts (e.g., signals driven by locus-specific technical features, dispersion-dominated associations without consistent burden direction, or regions prone to mapping ambiguity such as segmental duplications or olfactory-receptor clusters) were excluded from the primary Results counts (Artifacts: N = 6) but are reported in Supplementary Table S6.

### Epigenomic enrichment analysis

We assessed whether GWAS and fine-mapped variants were localized to regulatory elements active in human HSPCs using ATAC-seq datasets, aged HSPC multiome data, and clonal HSPC resources.

We tested meta-analysis lead SNPs for overlap with CD34⁺/HSPC ATAC peaks from the GSE305370 dataset. Lead variants were intersected with the peak set and then compared to the chromosome and minor-allele-frequency matched null variants drawn from the meta-analysis variant set, excluding genome-wide significant variants and variants within 500 kb of any lead. Using a fixed number of permutations, we computed peak-overlap fractions for observed and null sets and summarized enrichment by fold change over null and empirical P-values.

We then projected the observed and matched-null variants into an aged bone-marrow/HSPC multiome reference^39^ and assigned broad HSPC states (stem-like, erythroid, megakaryocyte, myeloid, lymphoid, cycling) based on predefined gene programs. For each of those states, we mapped variants to ATAC peaks and estimated enrichment for all, novel, and known lead sets by comparing state-specific peak overlap in observed versus null variants. In order to refine this analysis, we constructed metacells in the aged HSPC multiome using SEACells^48^ with a primary configuration (fixed metacell count, variable peak number, principal components, and random seed) and ran sensitivity analyses over alternative metacell counts, variable peak sets, seeds, and clustering algorithms. For each metacell configuration, we quantified enrichment for meta-analysis lead variants (directly and within defined genomic windows), a balanced genome-wide significant locus set, and fine-mapped variants of UKB at several posterior inclusion probability thresholds, using both unweighted and PIP-weighted summaries.

For human HSPC/HSC epigenomic enrichment, we focused only on fine-mapped variants from UKB with posterior inclusion probability being at least 0.2. We assembled a human HSPC/HSC peak panel from local multiome inputs and GSE305370 CD34⁺ resources and then compared observed variants with chromosome and MAF-matched null variants sampled from UK Biobank summary statistics, excluding variants near observed signals and requiring null variants to be non-significant under the GWAS model. For each resource, we computed direct peak overlap and continuous proximity/accessibility metrics based on nearest peak accessibility and distance, and then averaged these metrics across observed variants with posterior-probability weighting. These values were compared against matched-null distributions to obtain enrichment estimates and state-level z-scores, which were used to classify loci by HSPC state specificity and summarized in locus-level tables for figure generation.

To prioritize regulatory loci, we combined human HSPC peak overlap and proximity metrics, motif-disruption scores from position weight matrices, transcription-factor binding features from AlphaGenome, transcriptome-wide association signals, cytoband labels, and empirical-Bayes gene rankings for fine-mapped variants. Variants were grouped into loci, annotation scores were aggregated, and locus-level regulatory scores and ranks were computed and used in downstream visualization and interpretation.

We integrated clonal HSPC information from ReDeeM^39^ by selecting candidate genes and variant-linked peaks from the state-specificity and regulatory-prioritization analyses and summarizing gene expression and peak accessibility by age, HSPC state, clone bins, and clone identities in the ReDeeM single-cell data. These summaries were combined with the human HSPC epigenomic enrichment and state-specificity outputs to define germline epigenomic context groups and modules.

In parallel, we integrated AlphaGenome transcription-factor binding deltas with GWAS credible-set effect directions by aligning fine-mapping Z-scores to predicted changes in TF binding, and summarized these TF-track disruptions and their directions at the locus level. These outputs were used as additional annotation layers in the regulatory prioritization framework but did not alter the core GWAS, fine-mapping, or enrichment workflows.

### Phenome-wide association of germline CH PRS and somatic passenger burden

We performed a targeted phenome-wide association study with the European-ancestry cohort in the All of Us Research Program to evaluate associations between 31 age-related phenotypes and (i) a germline polygenic risk score (PRS)^49^ for clonal hematopoiesis and (ii) observed somatic passenger mutation burden with electronic health record (EHR) –derived phenotypes. Analyses were restricted to individuals in the EUR genetic cluster with whole-genome sequencing, complete covariate information, and available EHR follow-up information. All models used standardized exposures so that the odds ratios and hazard ratios correspond to a 1 standard-deviation increase in the germline PRS or residualized passenger mutation burden.

Germline CH PRS weights were derived using PRSFNN^49^, a neural empirical Bayes polygenic risk score method, trained on UK Biobank WGS SNP genotypes. Scores were then applied to All of Us EUR participants using plink2, followed by within-cohort standardization to mean 0 and variance 1. We specified a targeted set of 31 EHR-derived phenotypes, including hematologic malignancies, solid tumors, cardiometabolic traits, and major chronic diseases, and defined case-control status from diagnosis codes using previously curated phenotype algorithms. For each phenotype, we fit logistic regression models with the germline PRS as the primary exposure and adjusted for genetic ancestry principal components, sex when appropriate, and natural spline terms for age. For time-to-event analyses, we fit Cox proportional hazards models with age as the time scale, the same germline exposure, and the same covariate structure. Survival analyses used age at sequencing as the entry time and excluded diagnoses occurring on or before that date to focus only on the incident post-draw events.

Observed somatic passenger mutation burden was derived from the residualized mutation-count phenotype used in the main GWAS, which had been previously adjusted for age, sex, and smoking. For the PheWAS, this residualized burden was standardized to mean 0 and variance 1 and treated as the exposure. We then applied the same set of targeted EHR phenotypes as for the PRS. Logistic models used the standardized burden as the exposure and included the same covariates as the PRS models, and Cox models again used age at sequencing as the time axis, excluded prevalent events at or before blood draw, and adjusted for genetic principal components, sex, and smoking. For rare outcomes such as AML and ALL, Cox models were fit using a pre-specified fallback strategy through which the models were first run with the full PC covariate set, if this specification produced non-finite estimates or convergence issues, we refit with a reduced covariate set including only the first four principal components plus sex and smoking, recording the covariate specification used for each endpoint. All logistic and survival models were run separately for the germline PRS and observed passenger mutation burden, and results were summarized as odds ratios or hazard ratios per 1 standard-deviation increase in the exposure with corresponding confidence intervals and P-values across the phenotype panel.

### Spatial analysis of geographic heterogeneity in passenger mutation burden

We performed a spatial regression analysis^50^ in All of Us to test whether regional socioeconomic context, air quality, or chemotherapy exposure explained observed geographic heterogeneity in passenger mutation burden on chromosome 21. The input phenotype was the Chromosome 21 residualized mutation-burden measure, which had been previously adjusted for age, sex, and smoking in individual-level quasi-Poisson models. For the spatial analysis, we further adjusted this phenotype for ancestry and sequencing quality by fitting a linear model with the 16 genetic principal components that was available in the controlled tier data on AoU, sequencing depth, and contamination as predictors.

Participants were included if they had a non-missing residualized phenotype, blood-derived DNA, available principal components and sequencing-quality covariates, and non-missing geographic information in the controlled-tier All of Us data. Blood DNA status and specimen dates were obtained from the All of Us biospecimen tables, and the earliest blood DNA collection timestamp was used as the blood-draw date for each participant. Geographic location was derived from the OMOP observation table using the same observation concept as in the original geographic notebook. In this cohort, the available field was masked at the three-digit ZIP code level (ZIP3), so we extracted the first three digits of the recorded ZIP field and used ZIP3 as the participant-level geographic unit.

Area-level analyses were conducted at the ZIP3 level. We aggregated the individual-level adjusted phenotype by computing the number of analytic participants, the mean of the residualized PC, its standard deviation, and the prevalence of pre-blood-draw chemotherapy exposure for each ZIP3. Chemotherapy exposure was defined from All of Us procedure-occurrence records by selecting curated chemotherapy-administration concepts from the procedure vocabulary; for each participant we created a binary indicator of at least one qualifying chemotherapy procedure on or before the blood DNA collection date, and ZIP3-level chemotherapy prevalence was calculated as the proportion of analytic participants with pre-blood-draw chemotherapy in that ZIP3. In order to avoid unstable estimates from very small cells, ZIP3 regions with fewer than 10 analytic participants were excluded from area-level models, leaving 471 ZIP3 areas for regression.

We derived two additional ZIP3-level covariates from external data. Socioeconomic context was summarized using 2022 American Community Survey median household income through which the median income and total population were obtained at the ZCTA5 level, ZCTAs were mapped to ZIP3 by their leading three digits, and ZIP3-level income was computed as the population-weighted mean of ZCTA5 median incomes within each ZIP3. Air quality was summarized using EPA Air Quality System daily data for 2023–2025. For each monitoring site and pollutant, we computed daily AQI values and took the daily maximum across pollutants; these maxima were averaged over 3 years to obtain a mean AQI per monitor, then spatially linked to ZCTAs and aggregated to ZIP3 using population weighting, with nearest-monitor imputation for ZCTAs lacking monitors. All ZIP3-level covariates (income, AQI, chemotherapy prevalence) were standardized before modelling.

The primary area-level outcome for regression was the ZIP3 mean of the adjusted phenotype. We first fit ordinary least squares (OLS) models at the ZIP3 level: (i) Mean residualize PC ∼ income, (ii) Mean residualize PC ∼ income + AQI, and (iii) Mean residualize PC ∼ income + AQI + chemotherapy prevalence. Residual spatial autocorrelation from these OLS models was assessed using Moran’s I under Queen contiguity weights constructed from ZIP3 polygons, which were generated by dissolving Census 2020 ZCTA polygons by their leading three digits. Because the resulting national ZIP3 contiguity graph was disconnected, formal spatial autoregressive models were restricted to the largest connected contiguity component, which comprised 119 ZIP3 areas representing 60,674 participants. Within this component we fit both a spatial lag model and a spatial error model, each including standardized income, AQI, and chemotherapy prevalence as predictors, and compared them to the full OLS model using Akaike information criterion (AIC). All spatial analyses treated ZIP3 regions as the unit of observation, so the spatial-model sample size (n = 119) refers to areas rather than individual participants.

## Supplementary Information

Supplementary figures are contained in Supplementary Material S1; supplementary tables are contained in Supplementary Material S2.

## Code Availability

https://github.com/weinstocklab/AofU_passenger_calling https://github.com/weinstocklab/aofu_CH_PRS_phewas https://github.com/weinstocklab/gwasplot https://github.com/weinstocklab/SuSIE_WDL https://github.com/weinstocklab/UKB_regenie_workflow

## Data Availability

UKB data were accessed under application number 454659. UK Biobank data are available to approved researchers through the UK Biobank Access Management System (https://www.ukbiobank.ac.uk). This study used data from the All of Us Research Program’s Controlled Tier Dataset (Version: cdrv8 - R9), available to authorized users on the Researcher Workbench.

## Acknowledgements

We gratefully acknowledge *All of Us* participants for their contributions, without whom this research would not have been possible. We also thank the National Institutes of Health’s All of Us Research Program for making available the participant data and cohort examined in this study. This research has been conducted using the UK Biobank Resource under Application Number 454659. We gratefully acknowledge funding from the National Heart Lung and Blood Institute (NHLBI: R01HL168894), to K.N.C. and J.S.W. We thank members of the Weinstock lab for helpful feedback.

## Ethics Declaration

UK Biobank was approved by the North West Multi-Centre Research Ethics Committee (Ref: 11/NW/0382). All participants provided written informed consent. All participants from All of Us program provided informed consent for use of their de-identified data and biospecimens for research. Analyses using All of Us data in this study were conducted under an approved All of Us Research Program project and complied with all program data use policies, including the Ethical Conduct of Research Policy and the All of Us Data Resource Code of Conduct.

## Notes

### Competing Interest Statement

The authors have declared no competing interest.

