## supplementary figures for "Multi-ancestry analysis of 791K whole genomes reveals the genetic, geographic, and phenotypic correlates of somatic passenger mutations in blood"

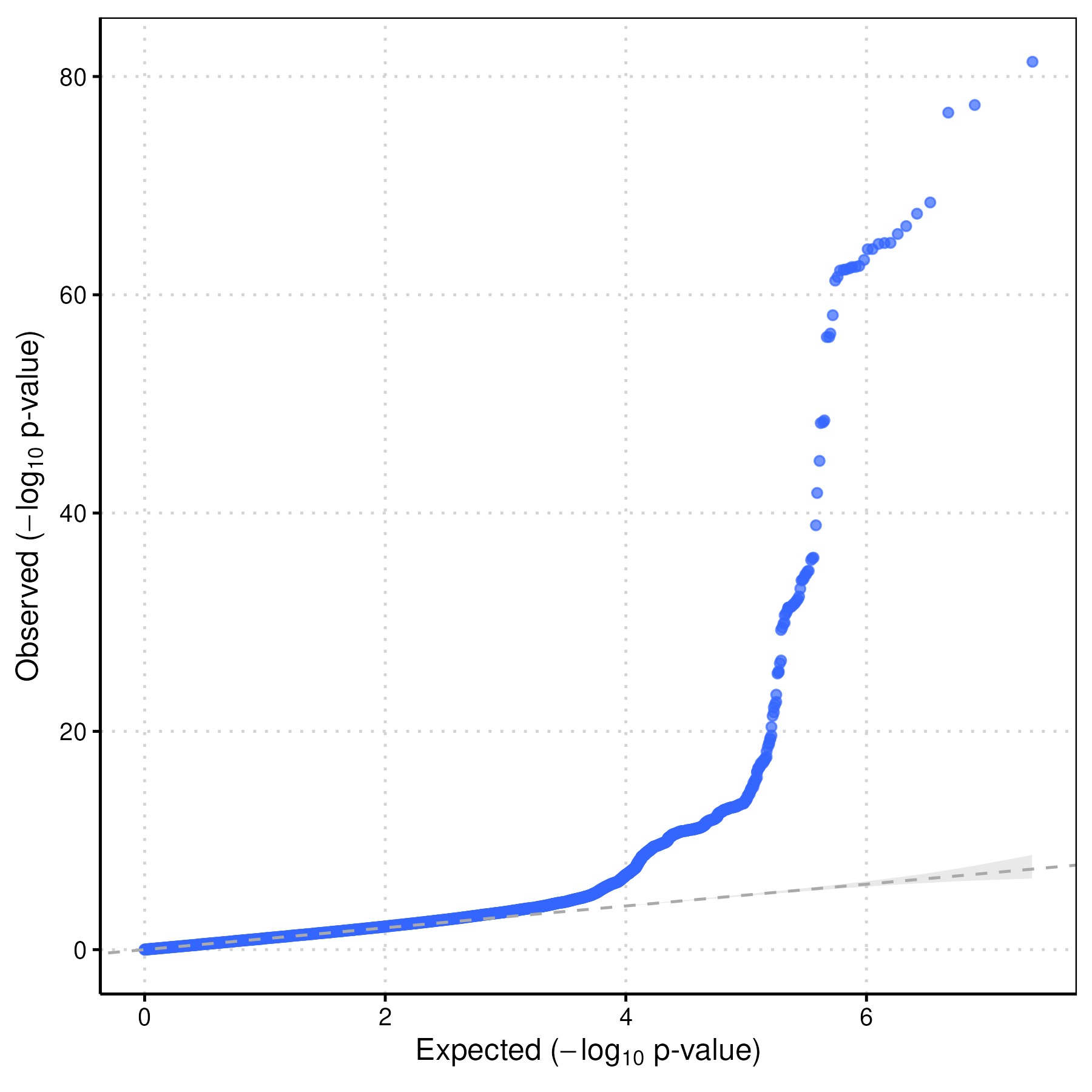

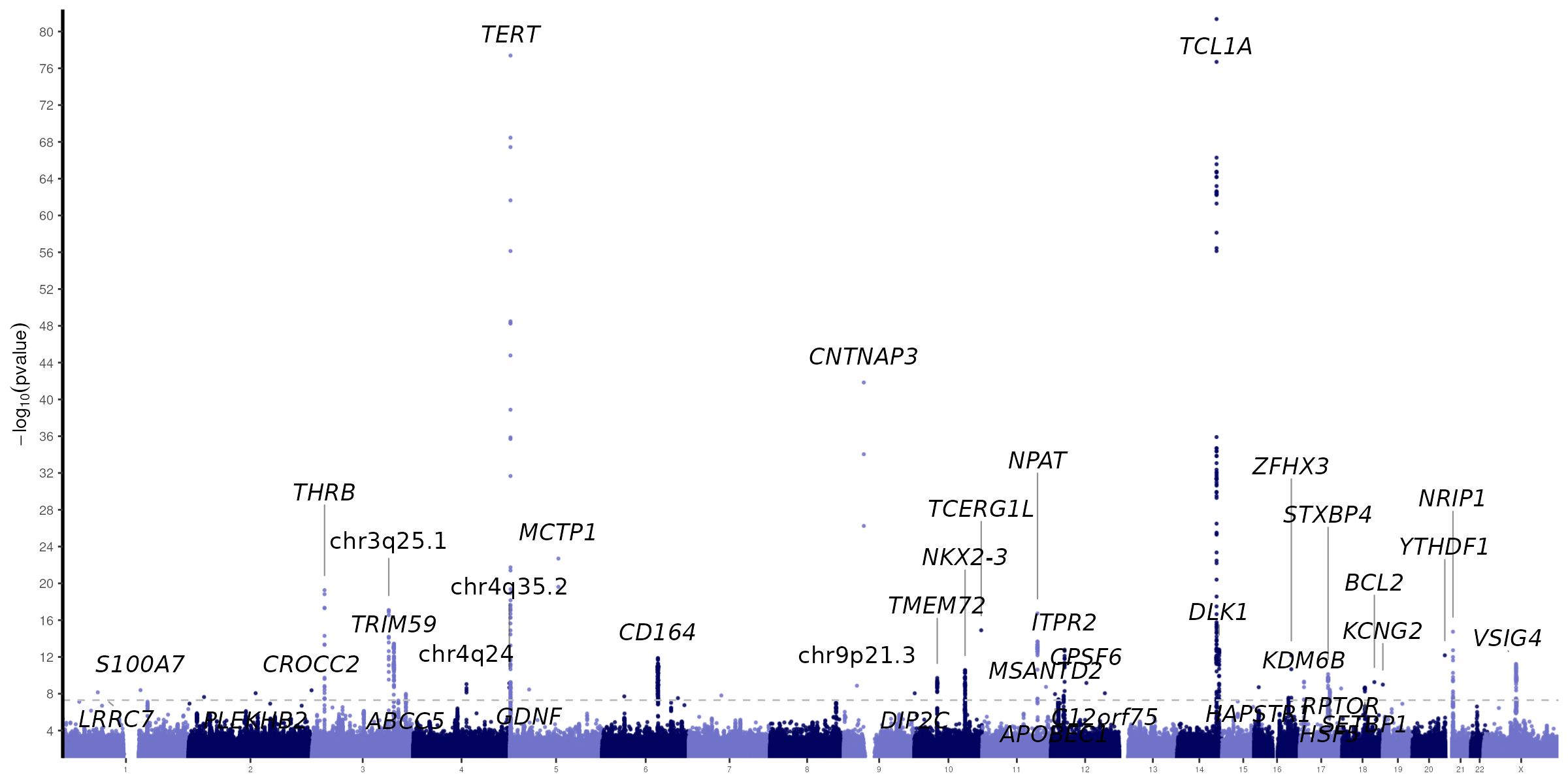

***Supplementary Figure S1*** *| Common-variant GWAS of somatic passenger mutation burden in UK Biobank. Quantile–quantile plot of observed versus expected −log10(P) values (top) and Manhattan plot of genome-wide association results (bottom) in 481,079 UK Biobank participants, identifying 45 genome-wide significant loci (P < 5 × 10⁻⁸); representative genes are labeled*

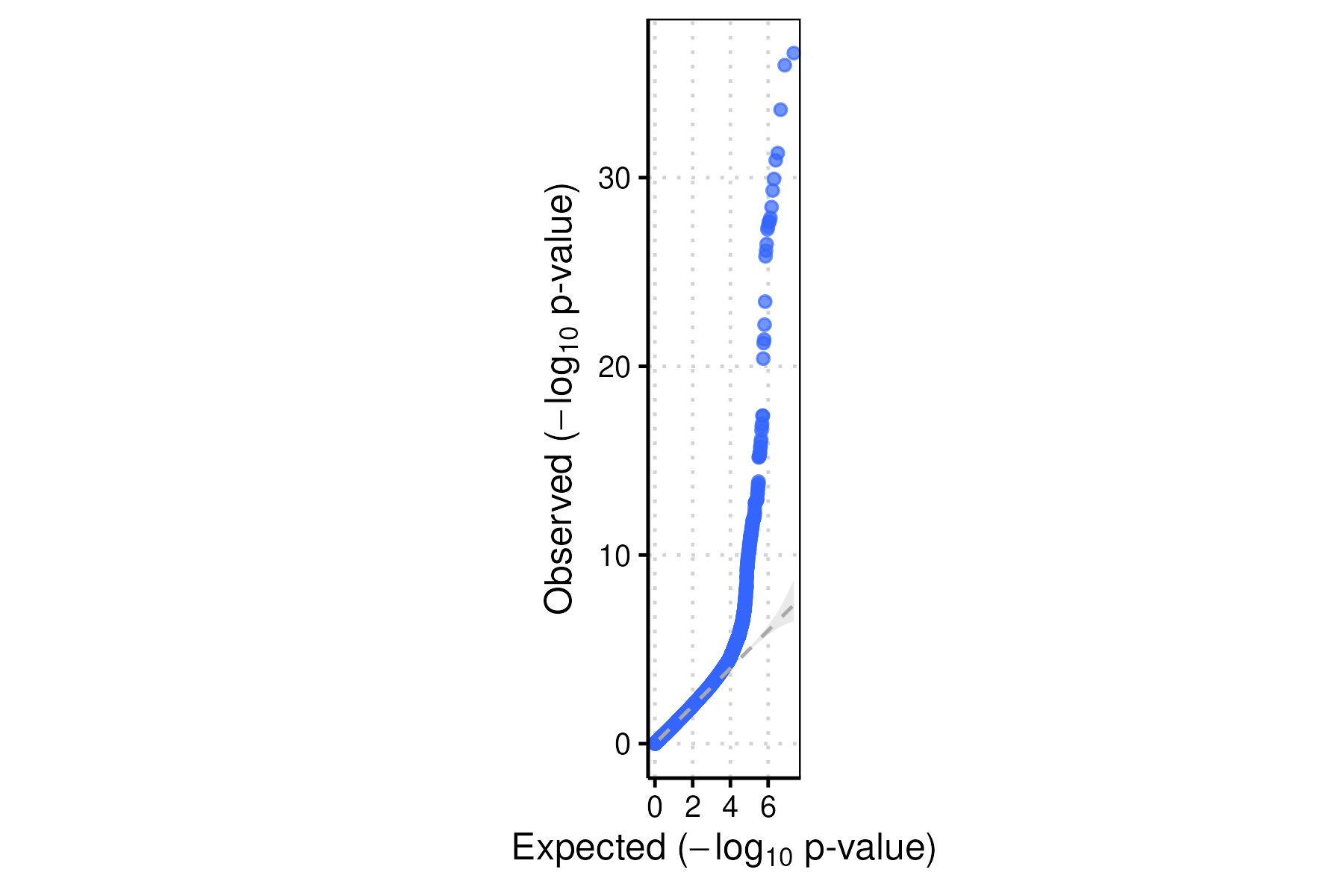

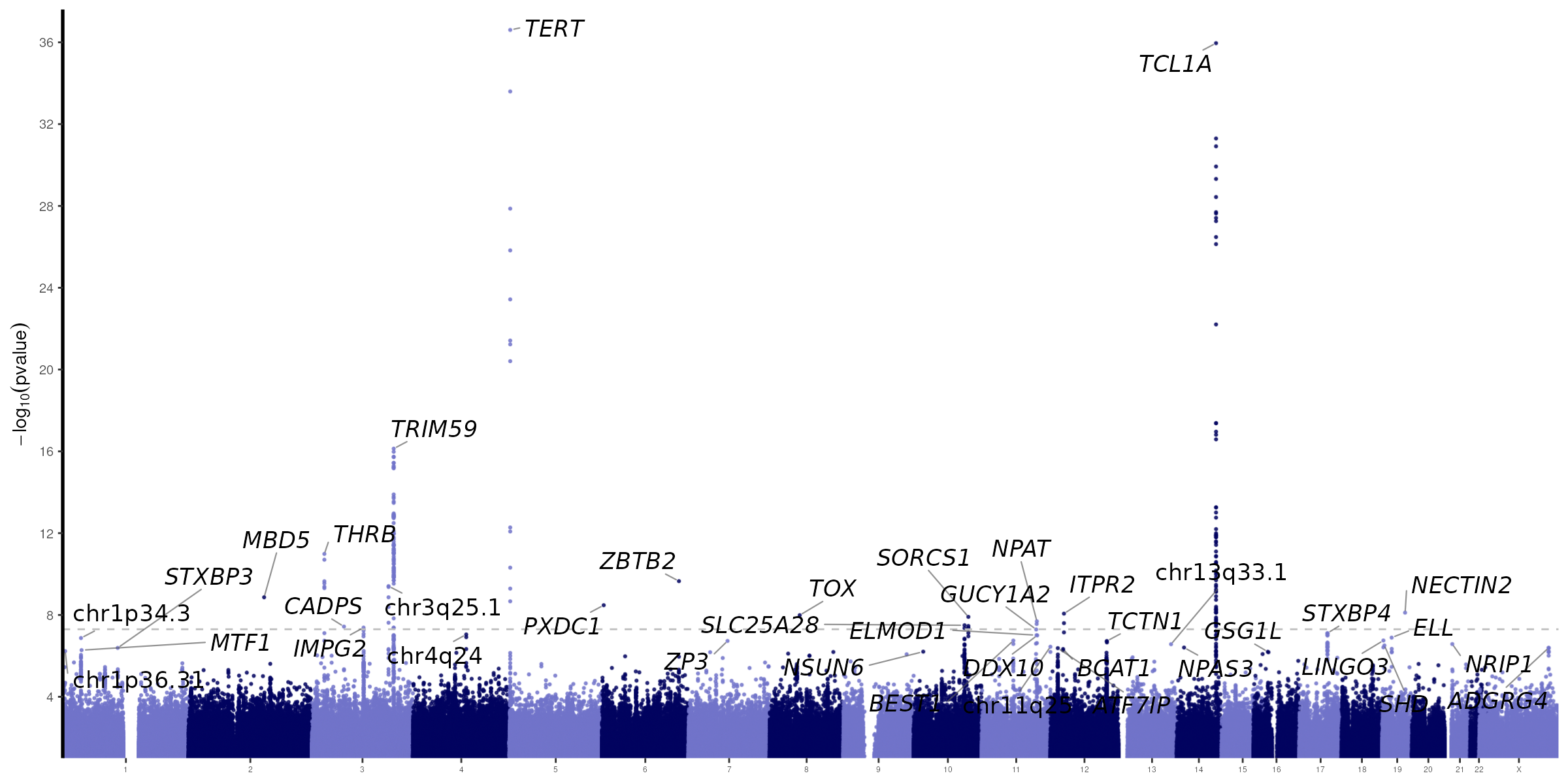

***Supplementary Figure S2*** *| Common-variant GWAS of somatic passenger mutation burden in All of Us. Quantile–quantile plot (top) and Manhattan plot of genome-wide association results (bottom) in 309,988 All of Us Research Program participants, identifying 16 genome-wide significant loci (P < 5 × 10⁻⁸); representative genes are labeled.*.
￼
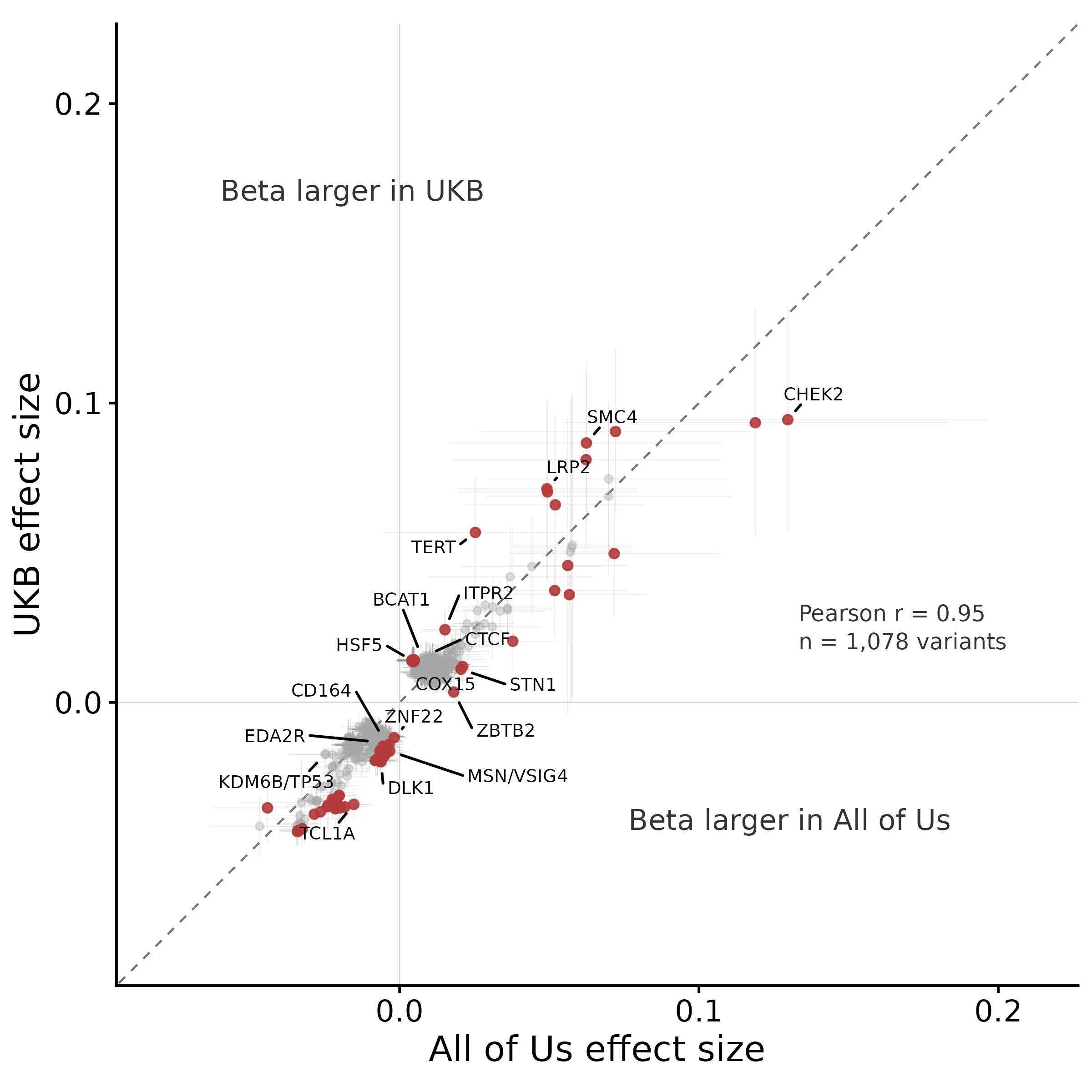

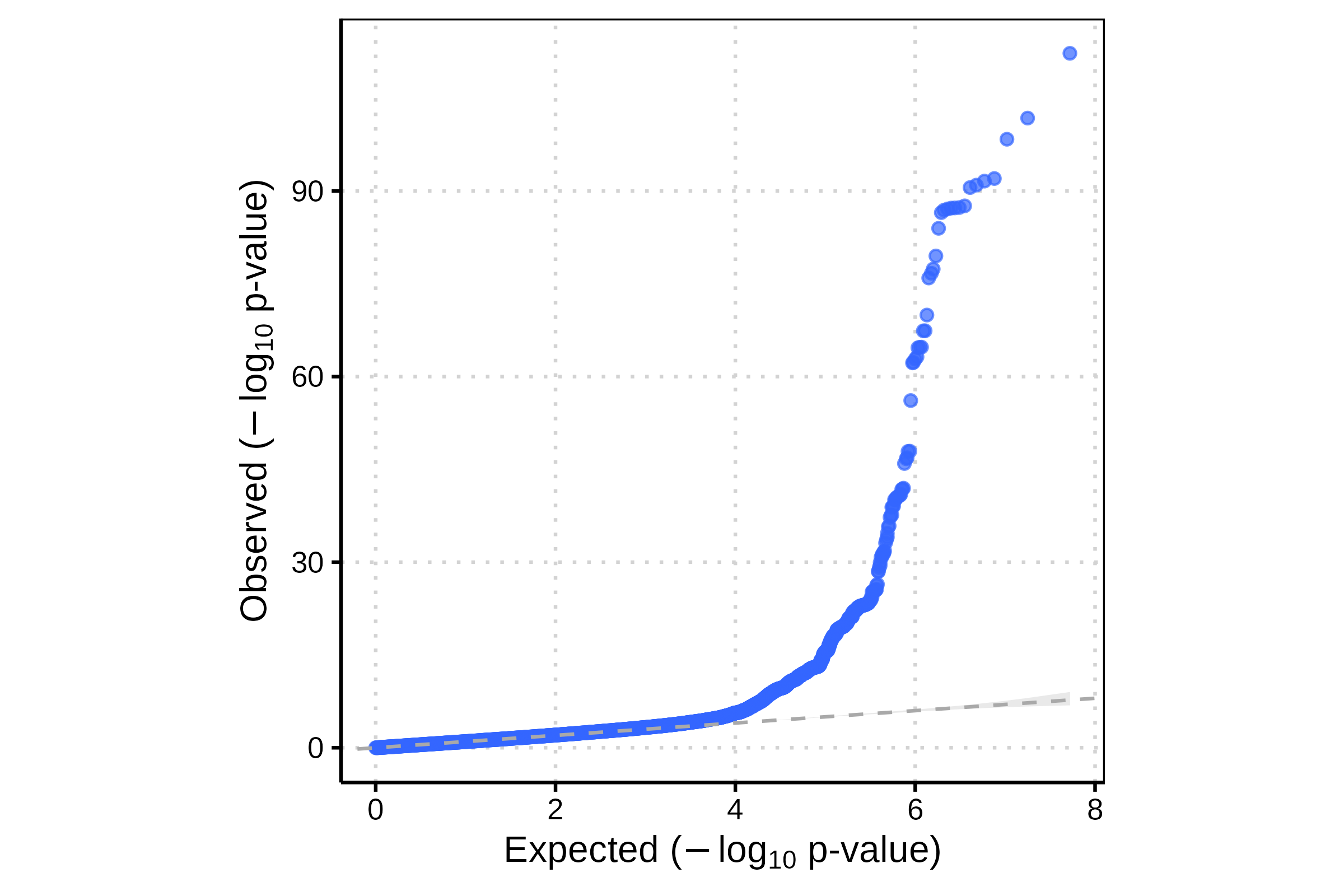

***Supplementary Figure S3*** *| Meta-analysis effect sizes and cross-cohort replication.* *Meta-analysis effect size (beta) plotted against minor allele frequency for the genome-wide significant loci identified in the UKB–All of Us meta-analysis (791,066 participants; 81 loci, P < 5 × 10⁻⁸) (top); novel loci are shown in red and previously reported/other significant SNPs in grey.*  *Replication of lead-variant effect sizes between cohorts: All of Us effect size versus UK Biobank effect size for n = 1,078 variants (Pearson r = 0.95); representative genes are labeled (middle). Quantile-Qunatile plot of the meta analysis (bottom)*

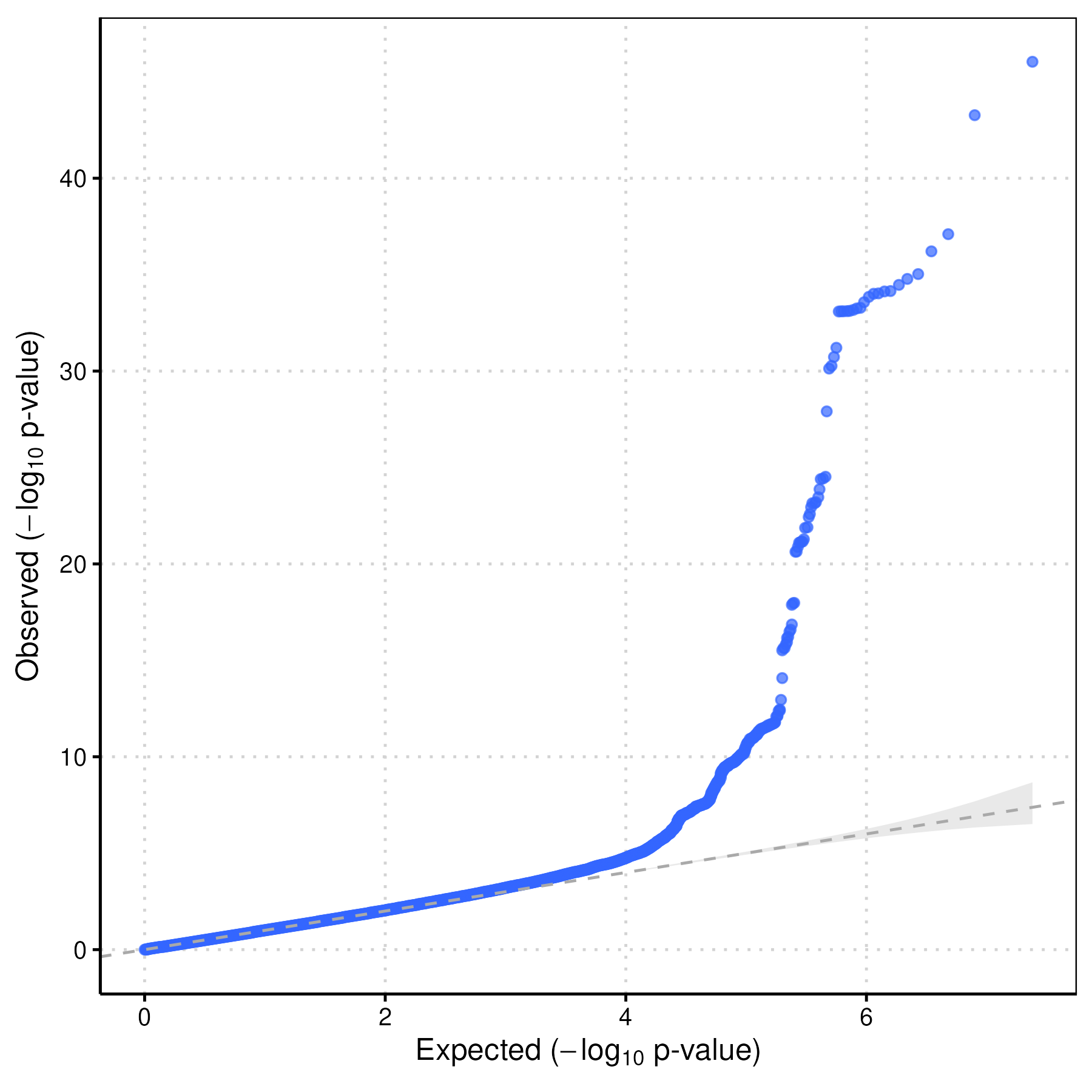

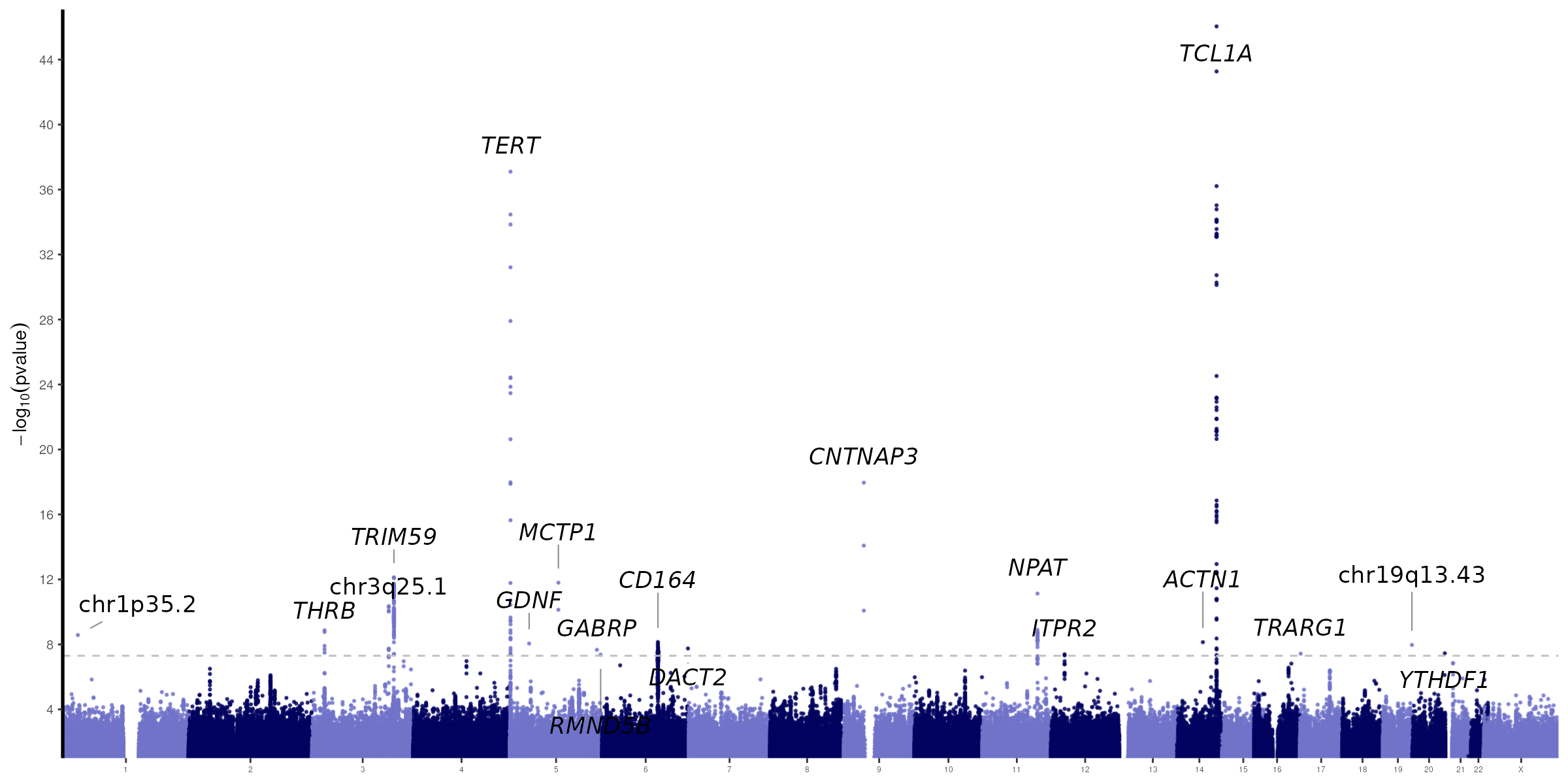

***Supplementary Figure S4*** *| Sex-stratified GWAS of somatic passenger mutation burden in females. Quantile–quantile plot and Manhattan plot of the female-only GWAS performed in UK Biobank; loci with female-specific or female-enriched effects (e.g., RMND5B, SMC4) are among those detected (see Figure 2 for comparison of male and female effect sizes at genome-wide significant loci).*

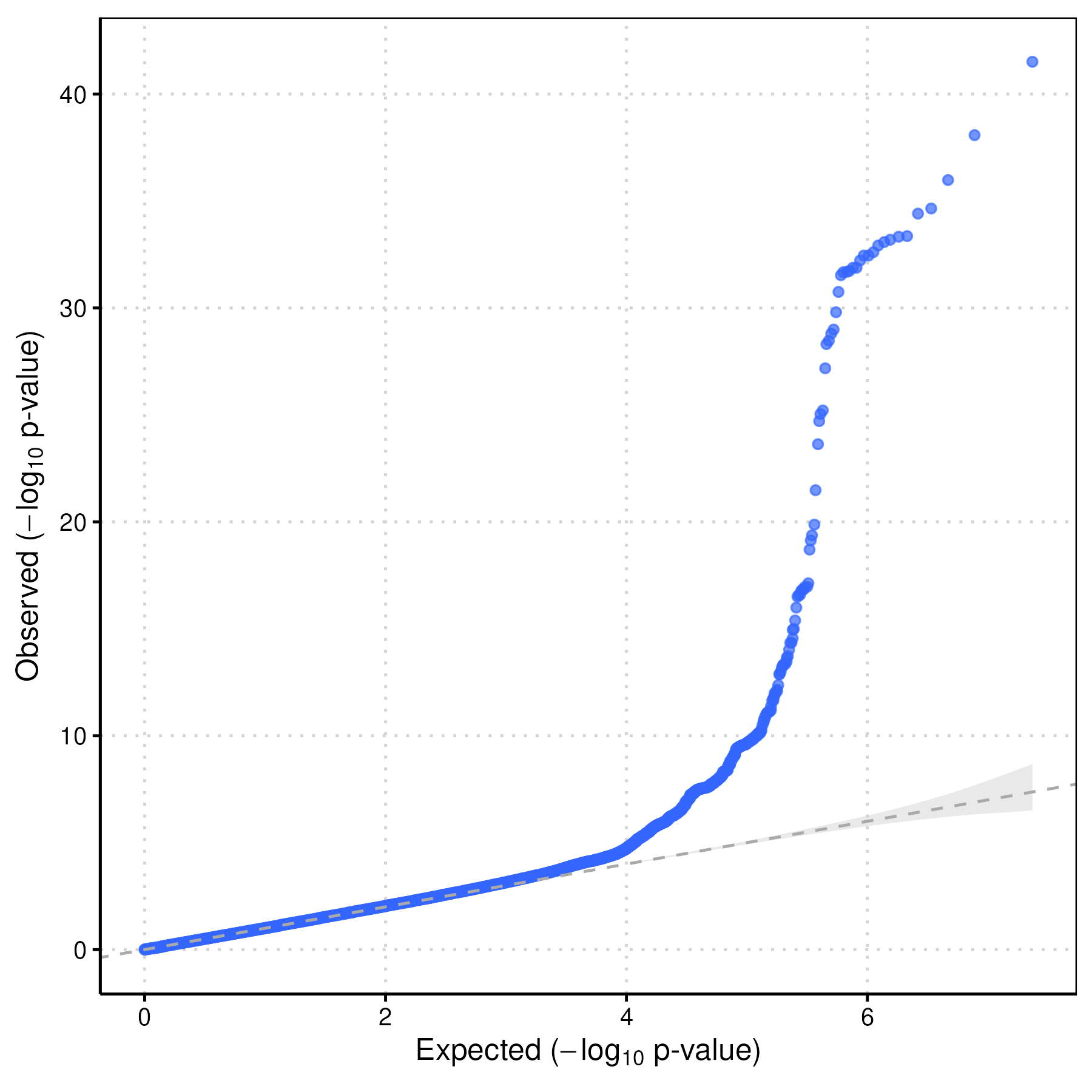

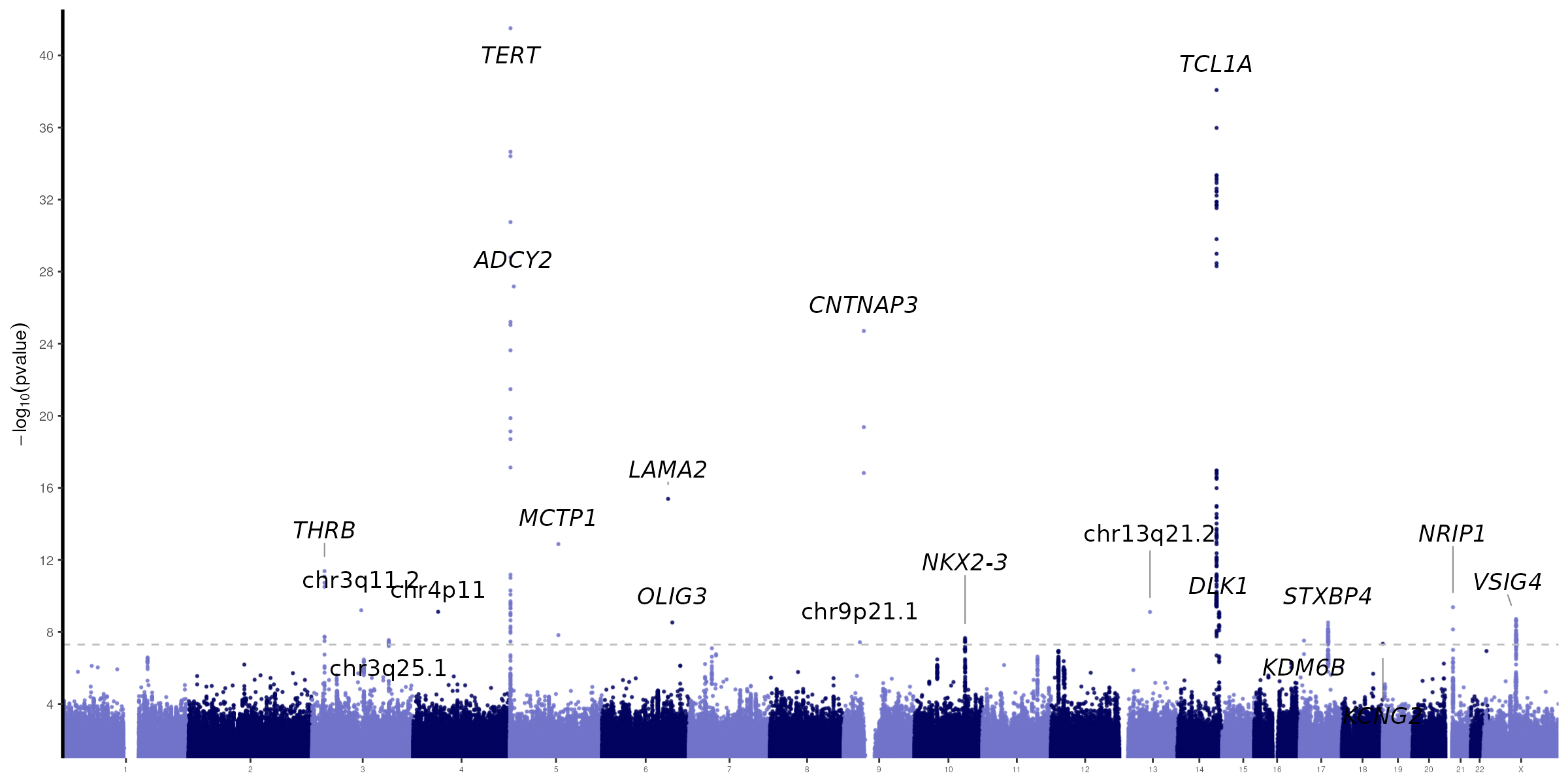

***Supplementary Figure S5:*** *Sex-stratified GWAS of somatic passenger mutation burden in males. Quantile–quantile plot and Manhattan plot of the males-only GWAS performed in UK Biobank; loci with male-enriched effects (e.g., OLIG3, ADCY2 ) are among those detected (see Figure 2 for comparison of male and female effect sizes at genome-wide significant loci).*

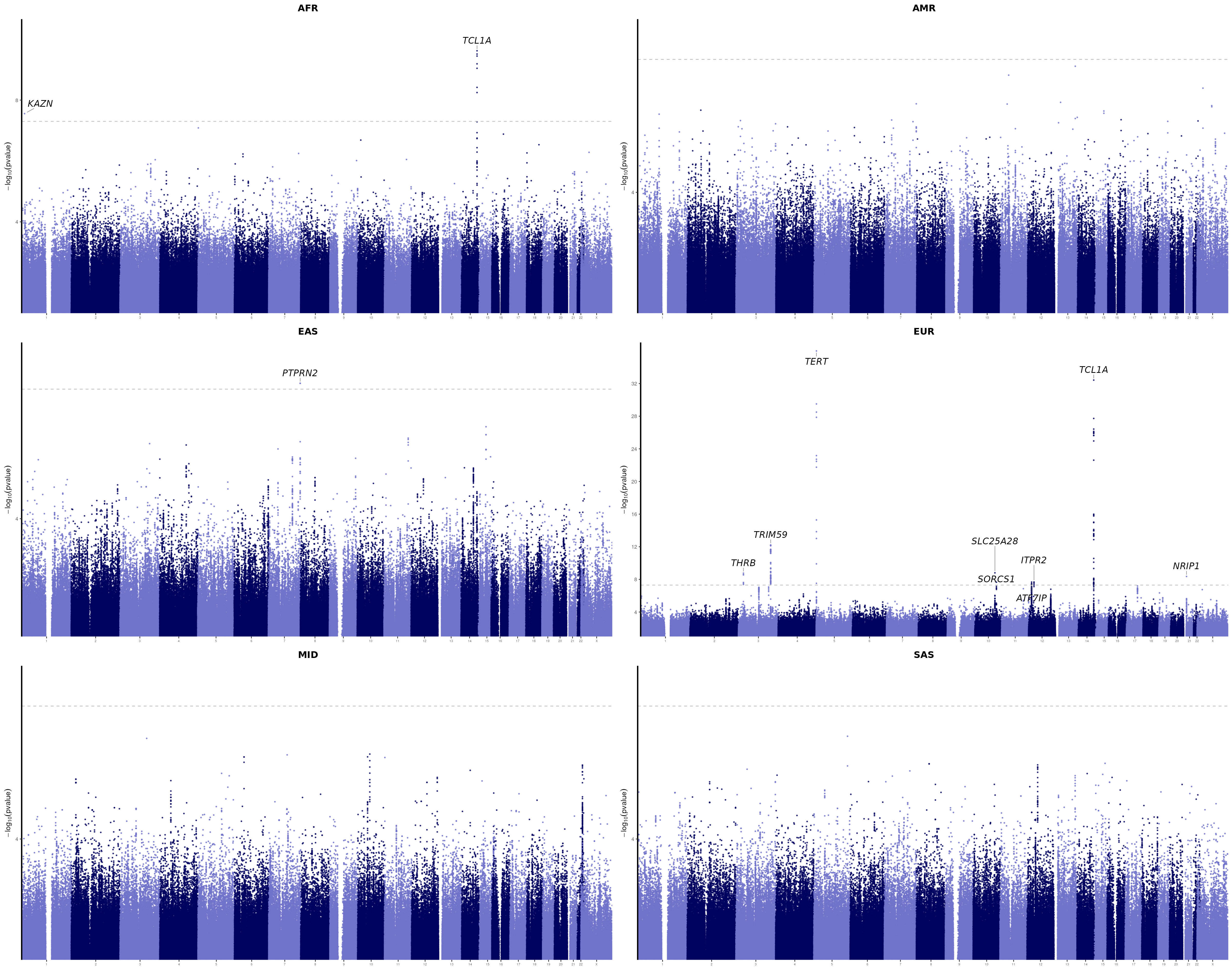

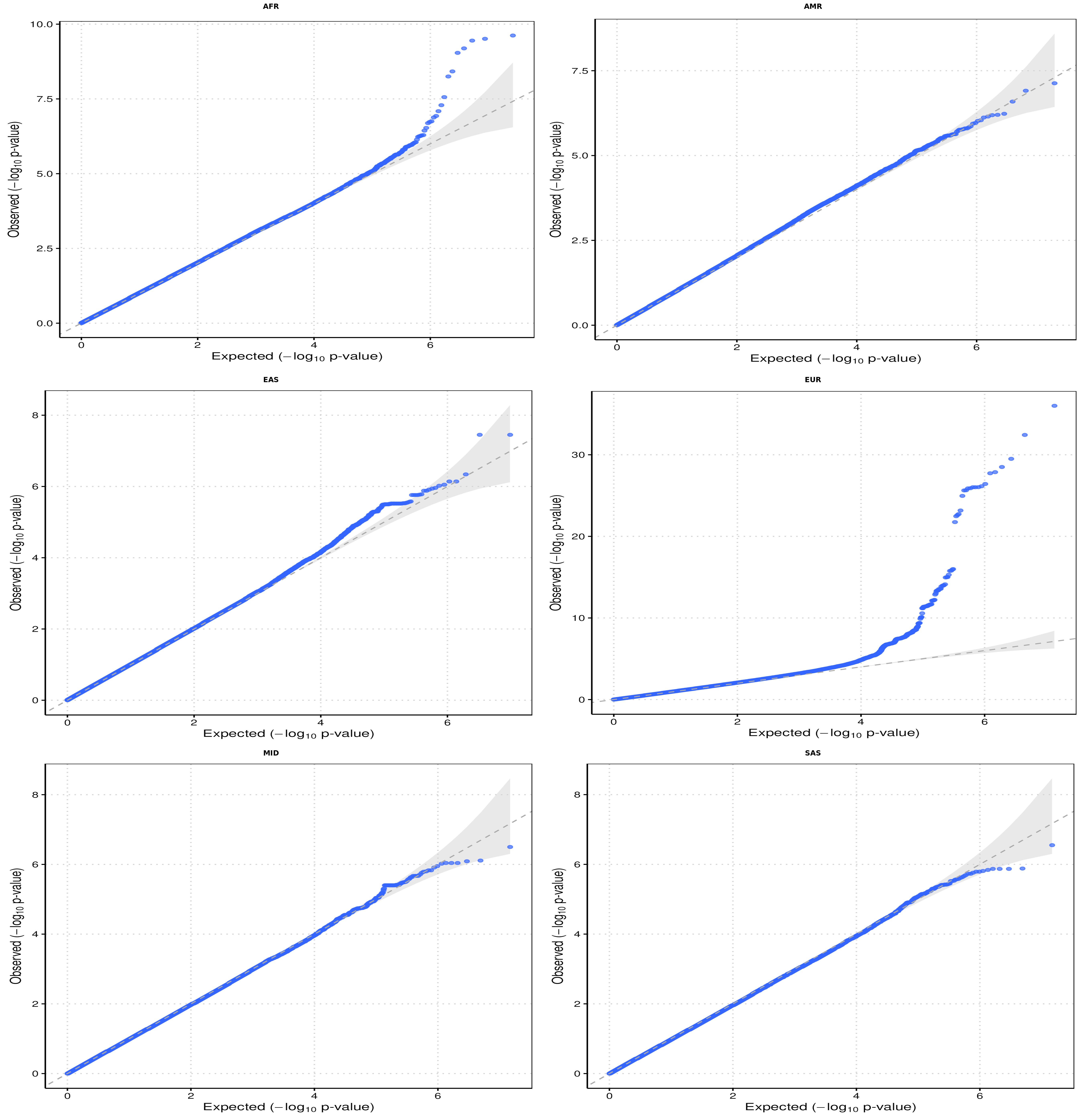

Supplementary Figure S6: Ancestry-stratified GWAS summary statistics in All of Us for AFR, AMR, EAS, EUR, MID, and SAS populations. Manhattan plots showing −log₁₀(p-value) across the genome for each ancestry group, with genome-wide significant loci (p < 5×10⁻⁸, dashed line) labeled by nearest gene. QQ plots comparing observed versus expected −log₁₀(p-values) for each ancestry group.

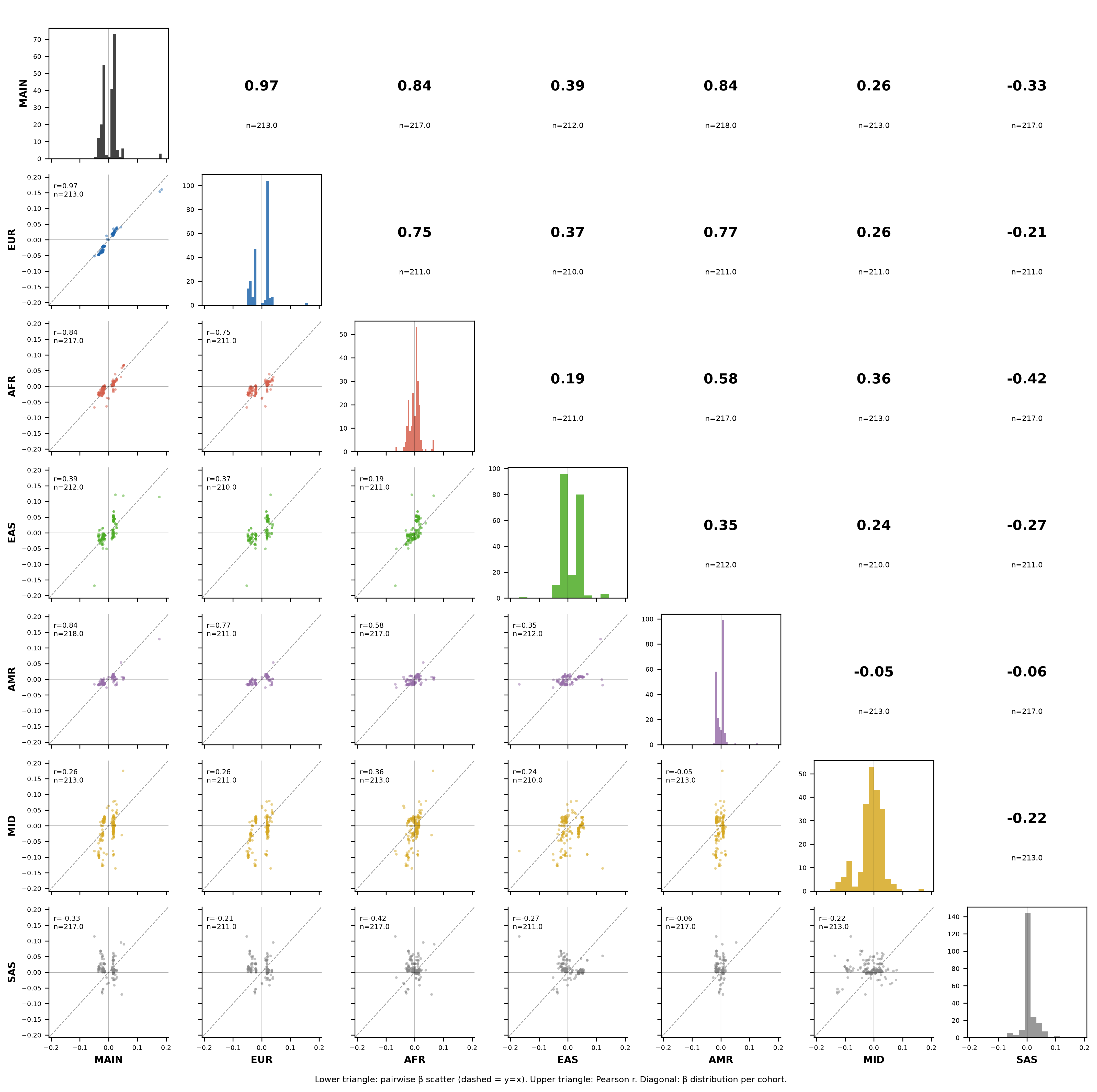

Supplementary Figure S7: Pairwise correlation of GWAS effect sizes across AoU ancestry cohorts. Effect sizes (β) for 220 SNPs reaching genome-wide significance (P < 5×10⁻⁸) in the overall AoU GWAS or in at least one ancestry-stratified cohort (EUR, AFR, EAS, AMR, MID, SAS). Diagonal panels show the β distribution per cohort; lower-triangle panels show pairwise β scatterplots, upper-triangle values show Pearson correlation coefficients with the number of SNPs in both cohorts.

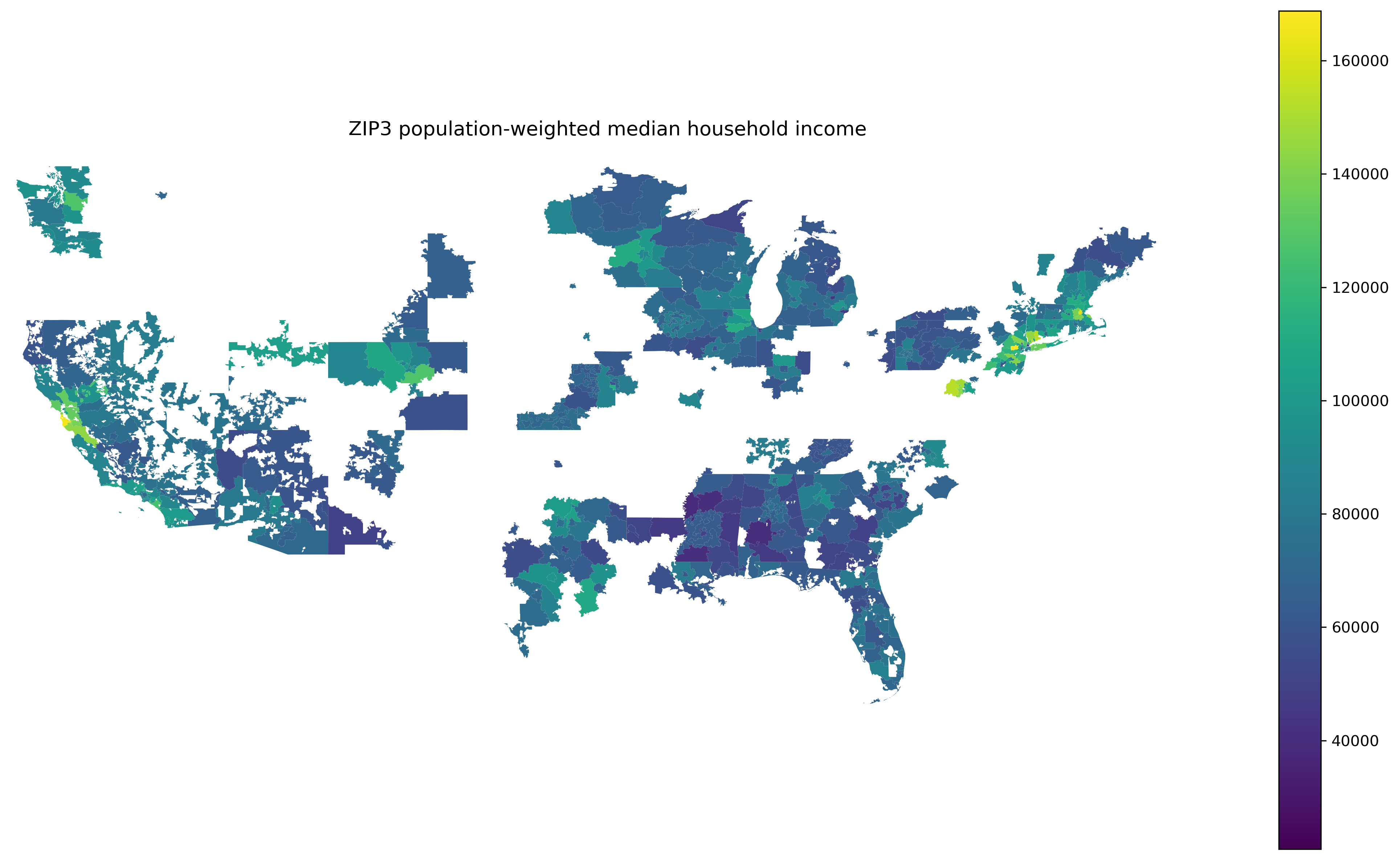

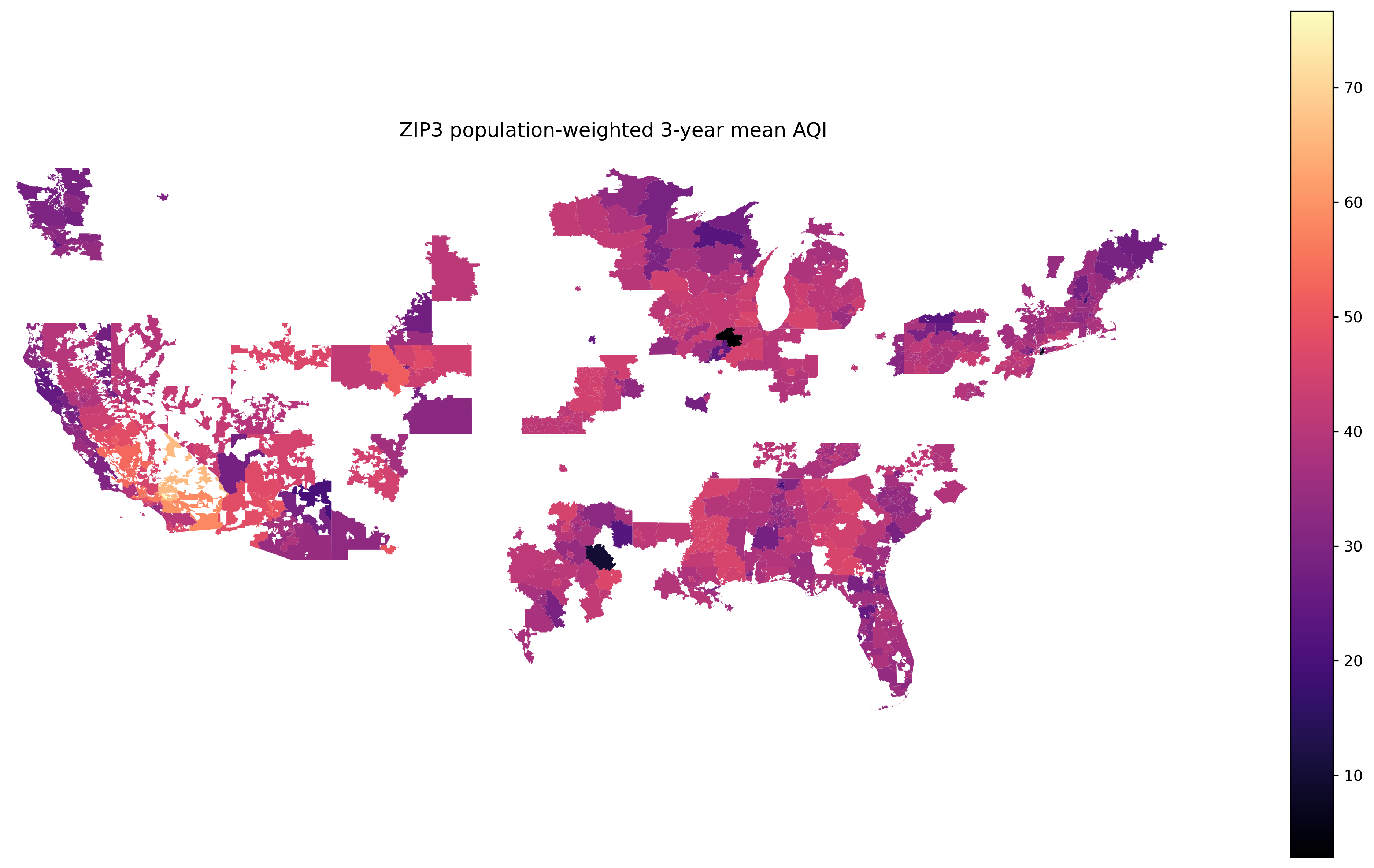

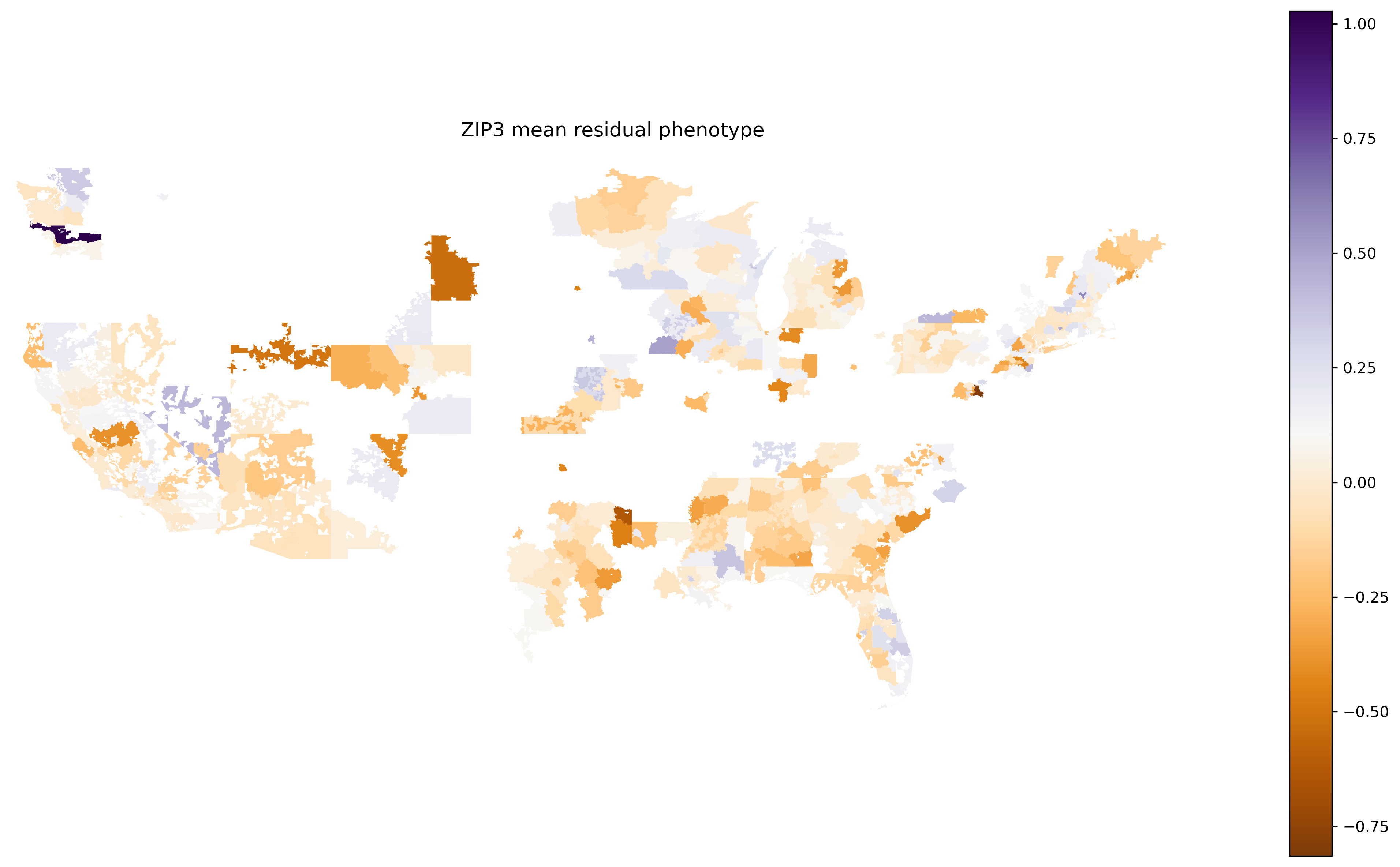

**S*upplementary Figure S8*** *| ZIP3-level covariates for spatial analysis of somatic passenger mutation burden. Maps show, across three-digit ZIP code (ZIP3) regions of the United States with ≥10 analytic All of Us participants (471 regions): population-weighted median household income (2022 American Community Survey); population-weighted 3-year mean Air Quality Index (EPA, 2023–2025); and mean residualized passenger mutation burden after adjustment for ancestry principal components and sequencing covariates. See Figure 7 and Methods for the corresponding spatial autoregressive modeling*
